# Predicting response to immune checkpoint blockade therapy among mismatch repair-deficient patients using mutational signatures

**DOI:** 10.1101/2024.01.19.24301236

**Authors:** Doga C. Gulhan, Vinay Viswanadham, Francesc Muyas, Hu Jin, Michael B Foote, Jake June-Koo Lee, David Barras, Youngsook L. Jung, Viktor Ljungstrom, Benoit Rousseau, Alon Galor, Bill H Diplas, Steven B Maron, James M. Cleary, Isidro Cortés-Ciriano, Peter J. Park

## Abstract

Despite the overall efficacy of immune checkpoint blockade (ICB) for mismatch repair deficiency (MMRD) across tumor types, a sizable fraction of patients with MMRD still do not respond to ICB. We performed mutational signature analysis of panel sequencing data (n = 95) from MMRD cases treated with ICB. We discover that T>C-rich single base substitution (SBS) signatures—SBS26 and SBS54 from the COSMIC Mutational Signatures catalog—identify MMRD patients with significantly shorter overall survival. Tumors with a high burden of SBS26 show over-expression and enriched mutations of genes involved in double-strand break repair and other DNA repair pathways. They also display chromosomal instability (CIN), likely related to replication fork instability, leading to copy number losses that trigger immune evasion. SBS54 is associated with transcriptional activity and not with CIN, defining a distinct subtype. Consistently, cancer cell lines with a high burden of SBS26 and SBS54 are sensitive to treatments targeting pathways related to their proposed etiology. Together, our analysis offers an explanation for the heterogeneous responses to ICB among MMRD patients and supports an SBS signature-based predictor as a prognostic biomarker for differential ICB response.

## INTRODUCTION

MMRD arises as a result of the inactivation of mismatch repair genes through germline or somatic mutation, promoter hypermethylation, or post-transcriptional gene silencing^1–6^. Genomes with MMRD are typically characterized by hypermutation, especially at microsatellite loci, due to polymerase slippage events, referred to as microsatellite instability (MSI)^7^. The high mutational burden fuels the production of neoantigens which are recognized by the adaptive immune system^8^, and the high neoantigen burden and immune infiltration levels make MMRD tumors immunogenic and thus suitable for ICB treatment.

However, a substantial fraction of MMRD patients does not respond to ICB treatment (30-60% for anti-PD-1/PD-L1 monotherapy^9–12)^. Previous studies identified the lack of a high burden of clonal mutations^13–16^ and truncating mutations in genes essential to mounting an anti-tumor immune response^17–21^ as negative genomic predictors of ICB outcome for both mismatch repair proficient (MMRP) and MMRD cohorts. However, these factors do not explain the difference in ICB outcomes observed *within* the better-responding MMRD group.

We hypothesized that mutational signatures, which are combinations of mutation types and their nucleotide contexts, may reveal the distinct mutational processes that underlie the variable ICB outcomes within MMRD tumors. Several signatures have already been associated with MMRD^22–26^, but no predictive model has been derived for the within-MMRD variation. Our results will show that, whereas MSI scores and mutational burden fail to distinguish the responders from non-responders, our signature-based approach does. Because much of the genomic profiling in available data has been performed on gene panels, mutational signatures sometimes must be inferred from a very small of mutations. We overcome this problem by utilizing the SigMA framework we developed^27^ and validated^28,29^ previously for detecting homologous recombination deficiency (HRD)^27^ from panel sequencing data.

## RESULTS

### MMRD classification combining SBS signatures and an MSI score

We analyzed a published^30^ cohort of patients (Supplementary Table 1) treated with PD-1/PD-L1 (n= 1307), CTLA-4-inhibitors (n = 99) or a combination thereof (n = 256) and profiled on gene panels. To identify MMRD cases, we developed a multivariate classifier (Supplementary Figure 1a) using features related to SBS signatures and microsatellite instability to classify tumors as MMRD, MMRP, or POLE exonuclease domain-mutant (POLE-exo). We validated our algorithm against MMR gene immunohistochemistry and MSI status computed using MSISensor^31^ for a subset of cases as well as for an additional cohort of patients who were not treated with ICB^32–34^ (Online Methods; Supplementary Figure 1b, Supplementary Table 2). Our MMRD prediction algorithm is applicable to gene panels with as few as 80 genes (∼0.2 Mb) (Supplementary Figure 1c) and outperforms methods based on microsatellite instability alone (Supplementary Figure 1c). Of a total of 1662 ICB-treated patients, 55 were classified as MMRD (PD-1/PD-L1, 52; CTLA-4, 1; Combo, 2), 4 as POLE-exo, and none as both (Supplementary Figure 2a). As expected, MMRD patients showed increased overall survival (OS) compared to MMRP patients (Figure 1a). When the tumor types were restricted to colorectal and esophagogastric cancers, which constitute the majority (43/55, 78%; Supplementary Figure 2a) of samples in our cohort, the difference between the MMRD and MMRP groups was even more pronounced (Supplementary Figure 2b).

**Figure 1.**
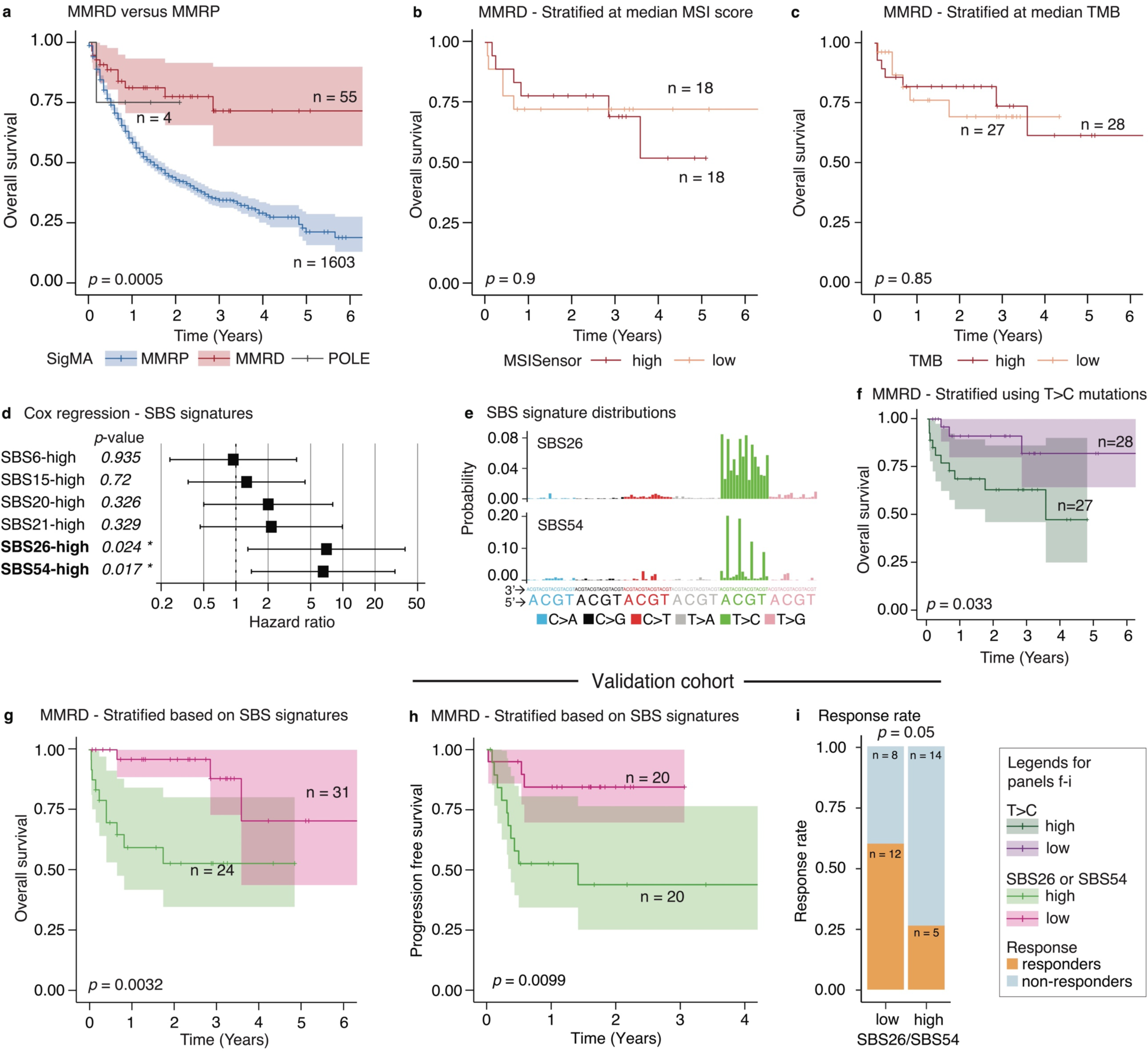
SBS signatures are predictive of ICB outcome. **a.** Kaplan-Meier (KM) curves of overall survival for MMRD, MMRP and POLE-exo mutant tumors; the *p*-value is for the comparison of MMRD versus MMRP groups. In panels b-i, we investigate further stratifications specific to MMRD patients. **b.** MMRD tumors stratified based on the median MSISensor score (not all cases had MSISensor scores available). **c.** Same as panel b, but for TMB. **d.** Hazard ratios for a Cox-regression model. Covariates are binary values defining whether the exposure of signatures are high (top quartile). Likelihood-ratio-tests are used for the hazard ratios. **e.** The distribution of trinucleotide frequencies for the SBS26 and SBS54. **f.** Same as panels b and c but stratified on the median T>C fraction. **g.** Based on the regression model (panel d), the SBS26-high or SBS54-high groups are compared to the double negative group. **h.** The KM curve of progression free survival (PFS) in the validation cohort for the signature-based stratification (no overall survival data were available for the validation cohort). **i.** Response rates of SBS26 or SBS54-high patients are compared. Complete and partial responders according to RECIST criteria were grouped into the responder category and the stable and progressive disease cases into the non-responder category (one patient in the validation cohort with unknown overall response but with PFS information was included in panel h but not j). For panels a-c, g, and h, the *p*-values for KM curves are obtained with the log-rank test; shaded areas indicate 95% CI.

### Two SBS signatures can stratify MMRD patients with differential ICB response

Next, we searched for genomic markers that could further stratify the 55 MMRD patients into responders *vs.* non-responders to ICB. We were motivated by the fact that tumor mutational burden (TMB) and a measure of microsatellite instability (MSISensor score) do not have a significant association with OS, whether stratified by the median (Figure 1b, c) or quartiles (Supplementary Figure 3). However, those in the lowest quartile of MSISensor scores among MMRD tumors tend to have shorter OS. Because almost all MMRD tumors already have a high TMB, TMB does not appear to have additional discriminating power, consistent with a previous study^35^.

Instead, we hypothesized that mutational signatures might provide more sensitive biomarkers to identify ICB responders among MMRD patients showing comparable TMB values. We built a Cox-regression model using binary covariates encoding whether a tumor carries a high exposure of each MMRD-related mutational signature detected in the cohort. This analysis yielded two signatures, SBS26 and SBS54, with significant hazard ratios: 7.03 (*p* = 0.024) and 6.55 (*p* = 0.017), respectively (Figure 1d), indicating a higher risk of death. We did not observe a tumor type dependence for these findings (Supplementary Figure 2d, f). Both signatures have a high probability of causing T>C mutations but distinct probabilities depending on their trinucleotide context (Figure 1e). Kaplan-Meier analysis confirmed that T>C-high tumors had a significantly shorter OS (Figure 1f) but also showed that the signature-based stratification gives a more refined identification of responders to ICB (*p* = 0.0032, Figure 1g). The groups identified by our signature-based approach do not differ in TMB or MSI scores (Supplementary Figure 2c), suggesting that signature analysis provides a measure distinct from other biomarkers that quantify the intensity of MSI^13^.

We validated our findings using three additional cohorts^35–37^ of MMRD gastric and colorectal cancer patients treated with PD-1 inhibitors (n = 40, Supplementary Table 3; although many other genomics studies on PD-1 inhibitor response are published, data are often not available publicly or clinical data were incomplete). Despite the small number of samples in our validation cohort, we found that progression-free survival was significantly shorter (Figure 1h, *p* < 0.01) for the SBS26-or SBS54-high (SBS26/54-high) samples. Response rates (complete or partial) were also significantly lower in SBS26/54-high samples (Figure 1i, *p* < 0.05). Together, our analyses (Figure 1, Supplementary Figure 4) reveal that mutational signatures represent a promising biomarker for identifying responders to ICB among MMRD patients.

### SBS26-high tumors carry mutations in DSB repair and replication fork stability genes

To understand the mechanisms underlying the lack of response to ICB in SBS26/54-high tumors, we compared the gene expression levels between the high and low signature exposure cases among the MMRD tumors from TCGA (expression data were only available for a small subset of the ICB-treated samples; there were 367 TCGA MMRD samples after removing those with hypermutation due to polymerase proofreading deficiency; Supplementary Tables 4 and 5). Pathway analysis based on gene expression for the first signature, SBS26, is summarized in Figure 2a (Online Methods, Supplementary Tables 6-8). SBS26-high tumors have striking upregulation in the DNA repair and cell-cycle pathways, including Fanconi anemia, double-strand break (DSB), and excision repair processes (pathways are vertical lines shown in Figure 2a; they are grouped into categories). Among the down-regulated pathways are apoptosis and autophagy, as well as immune system and angiogenesis.

**Figure 2.**
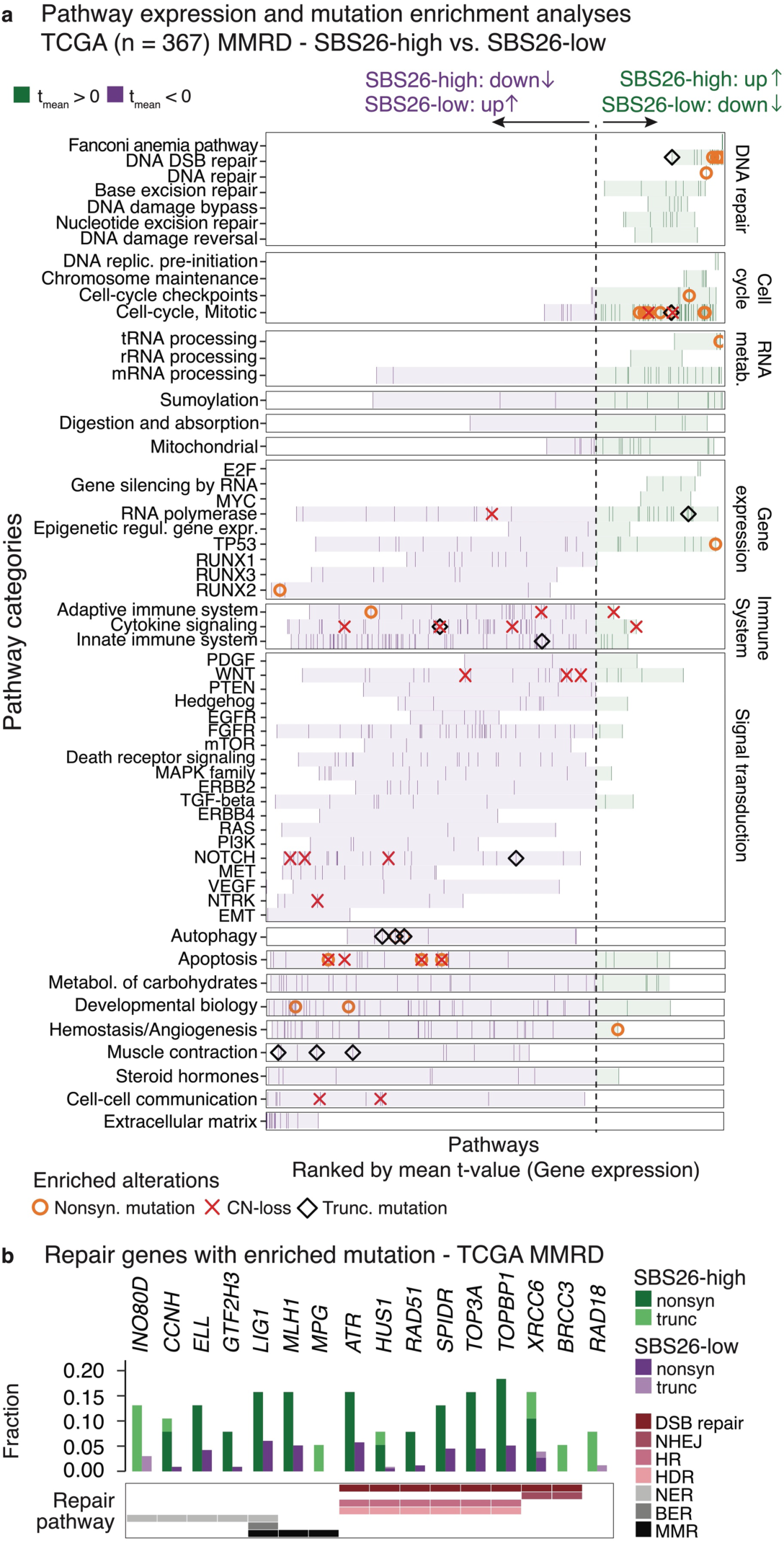
Pathway analysis reveals a connection between SBS26 and genomic instability. **a.** Pathways are ordered based on the rank of the t-value obtained from the comparison of gene set enrichment scores (Reactome^42^ and Hallmark^43^ gene sets obtained from MSigDB^44^) between SBS26-high and low TCGA samples. All pathways are shown, and the rank of each pathway is denoted as a vertical line (purple or green based on whether the t-value is > 0 or < 0). Pathways are grouped in each row according to their categories and subdivided into subcategories (for DNA repair, cell cycle, RNA metabolism, gene expression, immune system and signal transduction). The shading covers the range defined by the lowest and highest ranks in that (sub)category. Significant enrichments (*p* < 0.05) in mutation frequencies in pathways in SBS26-high tumors are also marked for different types of alterations (circle: nonsynonymous mutation, diamond: truncating mutation X: copy number loss). **b.** The fraction of samples with alteration on the DNA repair genes that have significant enrichment in the mutation frequency in SBS26-high tumors. The repair pathways are annotated below the barplot (NHEJ: Nonhomologous end joining, HR: Homologous recombination, HDR: Homology directed repair, NER: Nucleotide excision repair, MMR: Mismatch repair, BER: Base excision repair).

The enrichment of nonsynonymous and truncating mutations (orange/black marks in Figure 2a; Supplementary Figure 5) in DSB repair and various other DNA repair-associated genes underlies the upregulation of DNA repair pathways in SBS26-high tumors. Genes in repair pathways with a significant enrichment are shown in Figure 2b. These include genes that play essential roles in replication fork processing and restart under replication stress (e.g., *ATR, HUS1, RAD51, TOP3A, TOPBP1,* and *SPIDR*^38–40);^ the resulting deficiency in replication fork processing/progression is the likely cause of the upregulation in the Fanconi anemia pathway. Similarly, the replication fork instability and inefficient DSB repair leading to an increased level of DSBs—as evidenced by the significant overexpression of *H2AX*, whose staining is often used as a marker of DSBs (t-test, *p* < 0.0001)—results in the upregulation of DSB repair pathways. Another consequence of dysregulated DSB repair is the elevated level of chromosomal instability (CIN) (Supplementary Tables 9, 11) and the downregulation of apoptotic pathways^41^. These findings point to a connection between the origin of SBS26 and replication fork instability/DSB repair deficiencies.

As an orthogonal validation for the characteristics of SBS26-high tumors, we examined the response of 51 MMRD cell lines to various drugs using data from the Genomics of Drug Sensitivity in Cancer (GDSC) project^45^ (Online Methods, Supplementary Table 12). We found that the drugs with the most significant increase in sensitivity in SBS26-high cell lines (Supplementary Figure 6a, Supplementary Table 13) were inhibitors of ATR (a key component of DSB repair and replication fork stability) and USP1 (which regulates Fanconi anemia pathway^46^). This is in line with the CIN and the upregulated Fanconi anemia pathway observed in the SBS26-high tumors in the TCGA data.

### Characterizing the chromosomal instability in SBS26-high tumors with rearrangement signatures

Mutational signature analysis using structural variants (SV)^47,48^ gives additional insights on the origin of CIN in SBS26-high MMRD tumors. We identified 11 SV signatures in the Pan-Cancer Analysis of Whole Genomes (PCAWG) consortium^49^ whole-genome sequencing pan-cancer data (Supplementary Figure 7), and compared the SBS26-high (n=12) *vs.* SBS26-low (n=26) MMRD cases. Two signatures, SV5 and SV6, representing small (<30kb) and medium (30kb-1Mb) deletions (Figure 3a), respectively, were significantly enriched in the SBS26-high compared to the MMRP tumors (Figure 3b, Supplementary Figure 8). SV5 has been shown to be prominent in tumors with HRD, especially those with bi-allelic *BRCA2* inactivation, *BRCA2*-/-(*p* < 0.0001, Figure 3c), and with Fanconi anemia repair deficiency (Supplementary Figure 8b)^50^. We find that SV6 is a distinct process and it is enriched in significantly delayed/under-replicated regions^51^ (Figure 3d). These observations confirm that CIN in SBS26-high tumors is likely due to intrinsic DSB repair deficiencies, at a lower intensity than in HRD tumors (average SV5: 94.6 and 15.4 in *BRCA2*-/- and SBS26-high MMRD, respectively), and replication fork instability.

**Figure 3.**
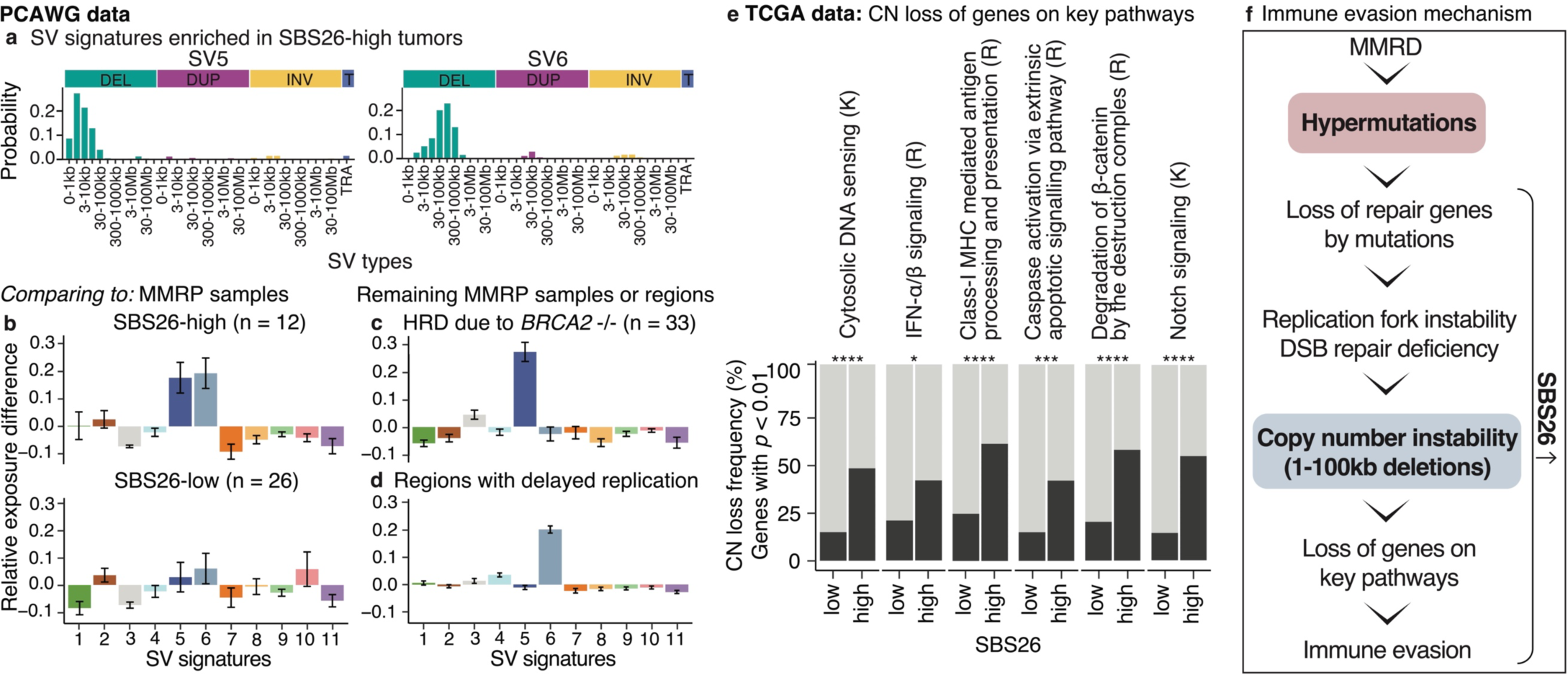
Small deletions in SBS26-high tumors lead to immune evasion. **a.** The probability distributions of the two SV signatures that are enriched in SBS26-high tumors (For the remaining signatures and extended SV types, see Supplementary Figure 7). **b.** The difference in the relative exposures of SV signatures between MMRD and MMRP samples in the same tumor types are calculated and averaged; shown for SBS26-high (top) and SBS26-low (bottom) samples. **(c)** Same as panel b, comparing MMRP, *BRCA2*-/-HRD samples to remaining MMRP samples **(d)** Same as panels b and c, but comparing SVs overlapping with fragile sites to all SVs, differently from panels b and c SVs in different regions of the same samples are compared and not different groups of samples. The error bars in panels (b-d) denote standard error. **e.** For key pathways, the fractions of samples with a CN loss on one of the genes with an enriched loss frequency (*p* < 0.01) are shown. K: Kegg, R: Reactome. Fishers exact test: *: < 0.05, **: 0.01-0.001, ***: 0.001-0.0001, ****: < 0.0001. **f.** A summary of key events leading into immune evasion.

One consequence of CIN in SBS26-high tumors is the frequent copy number (CN) losses of genes in anti-tumor immune response pathways (marked by “X” in Figure 2a), potentially explaining the worse outcome of this group to ICB. The pathways with differential CN losses include cytosolic DNA sensing, type-I interferon (IFN) signaling, and Class-I MHC antigen presentation pathways (Figure 3e). The reduction of class-I MHC antigen presentation by the loss of HLA alleles or *B2M* gene has been reported as a mechanism of resistance to ICB^52^. Cytosolic DNA sensing with the STING pathway has been shown to play a role in MMRD tumors through type-I IFN signaling^53,54^. In addition to the more frequent loss of *STING1* in SBS26-high samples (*p* = 0.027, Fisher’s exact test), we find that nearly half of the SBS26-high tumors are affected by CN losses that impair the DNA-sensing pathway. A similar situation holds for the IFN-α and β pathways. Others with enriched CN losses and potential connections to immune evasion were the Wnt and Notch pathways (Figure 2a, 3e). Specifically, loss of degradation of β-catenin by the destruction complex and degradation of DVL leading to a higher abundance of β-catenin may result in immune exclusion^55^. Loss of the Notch signaling pathway genes can also indirectly lead to upregulated Wnt signaling through β-catenin^56–58^. Because point mutations were not enriched in these pathways, we infer that CIN, introduced by SBSs and indels on DNA repair genes, leads to CN losses that trigger immune evasion (Figure 3f). A high burden of CN losses was previously reported to lead to unfavorable treatment outcomes in tumors treated with ICB^16,51,59^.

### SBS54-high tumors represent a distinct subtype linked with transcription

We repeated the pathway enrichment analyses for SBS54. SBS54-high tumors share some properties with SBS26-high tumors, such as rapid proliferation and upregulated DNA repair as well as over-expression of *MYC* (*p* = 0.017, t-test) and Myc targets, which can have immuno-suppressive effects^60,61^. But, upregulation of DNA repair pathways is less prominent, and the repair pathway with the highest enrichment is base excision repair rather than DSB repair. Instead of high CIN, SBS54-high tumors show strong upregulation of RNA-processing and RNA metabolism (Supplementary Figure 9), suggesting a transcription-related origin for SBS54. Consistent with this, SBS54 exposure is enriched in coding regions compared to non-coding regions (Supplementary Figure 10), and for SBS54-high cell lines, drugs targeting chromatin histone acetylation/methylation and RNA polymerase-1 had significantly higher sensitivity^62,63^ (Supplementary Figure 6b).

Note that we also sought to identify other immune-related characteristics^15,54,64,65^ that may be specific to the SBS26-high/SBS54-high TCGA tumors. These include immune cell proportions, TCR repertoire richness^66^, HLA loss-of-heterozygosity (LOH) frequency, neoantigen counts, and subclonal structure. Although there were some differences, they were not shared across tumor types (Supplementary Figure 11, 12). These results indicate that the signature-based analysis provides information about the MMRD tumors not captured by these parameters.

## DISCUSSION

In summary, we have shown that the classification of MMRD tumors using a mutational signature framework can distinguish responders from non-responders to ICB treatment. Signatures capture characteristics of MMRD tumors beyond current biomarkers, which quantify the intensity of MSI (Supplementary Figure 13) and tumor heterogeneity, and therefore, they can be used as a complementary measure. Our characterization of the tumors with either of the two signatures (SBS26 or SBS54) in the TCGA data and cancer cell lines revealed distinct properties of these tumors that explain the difference in the outcome, including dysregulation of DNA repair pathways and genome instability that leads to CN loss of immune signaling genes. Whereas standard signature analysis requires exome or genome sequencing, the proposed method can be applied to panel sequencing data despite a >100-fold reduction in the number of mutations detected on panels. Given the widespread adoption of gene panels in clinical settings, the proposed approach may serve as a useful complement to the detection of actionable gene alterations. The majority of the cohorts we examined consisted of colorectal and esophagogastric cancers; thus, our findings should be validated in other tumor types before the conclusions can be extended. Previous studies in pediatric cancers^26,67^ found that MMRD samples with POLE-exo mutations are marked with an indel^26^ and an SBS signature (SBS14)^23^ and have higher ICB response rates compared to MMRD alone. The prevalence of MMRD with POLE-exo in colorectal and esophagogastric cancers is substantially lower than in pediatric cancers; the cohorts in this study did not contain any such samples. The role of SBS14 should be further investigated in adult cancers. As the tumor microenvironment plays an essential role in anti-cancer immunity, future studies should consider combining the signature-based biomarkers with more refined variables that capture the key features of the tumor environment.

## ONLINE METHODS

### Datasets

#### TCGA data

Mutation calls from the whole exome sequencing (WES) data (curated through the use of multiple SNV callers ^68^) and gene expression matrix were obtained through the PanCanAtlas data portal https://gdc.cancer.gov/about-data/publications/pancanatlas (files: mc3.v0.2.8.PUBLIC.maf.gz, EBPlusPlusAdjustPANCAN_IlluminaHiSeq_RNASeqV2.geneExp.tsv). The gene level copy number alterations calculated by GISTIC^69^ are provided at https://gdac.broadinstitute.org/. Finally, MSISensor scores, TCR.Richess measures were obtained from a previous PanCanAtlas publication^66^ using the same data portal.

#### WGS data

SBS and SV calls from the Pan-Cancer Analysis of Whole Genomes (PCAWG and ICGC) study were downloaded from the ICGC data portal (https://dcc.icgc.org/). SBS calls were derived from three established mutation calling algorithms; SV calls by the PCAWG Structural Variation Working Group were identified by at least two of the four algorithms applied^44^.

#### MSK-IMPACT panel data

The nonsynonymous and truncating SBS calls for MSK-IMPACT panels used in the ICB survival analysis were downloaded from cbioportal (https://www.cbioportal.org/) of the related publication^30^. For a subset of samples, synonymous and intronic mutations were also available from an earlier publication^70^. For signature analysis and MMRD identification, all the mutations are informative, and thus also the synonymous and intronic mutations were used when available. The MMR (MSH2, MSH6, MLH1, and PMS2) immunohistochemistry results were obtained from the Supplementary Material and cBioPortal pages of publications that focus on colorectal and endometrial cancers^32,33^. MSISensor scores for colorectal, endometrial and esophagogastric cancers were obtained from previous studies ^32–34^.

#### Validation cohorts

The data from Rousseau, Foote *et al.* cohort^36^, which consisted of somatic mutation calls (Indel and SBS), OS, PFS, best overall response, MSISensor scores and TMB, was obtained through communication with authors after signing a data usage agreement.

The raw sequencing data from Kwon^37^ *et al.* and Bortolomeazzi^35^ *et al.* were accessed from European Nucleotide Archive (PRJEB40416) and Euorpean Genome-Phenome Archive (EGAD00001006165, EGAD00001006164), respectively. The raw data were mapped against the GRCh38 build of the human reference genome using BWA-MEM^71^ (0.7.17-r1188) Aligned reads in BAM format were processed following the Genome Analysis Toolkit^72^ (GATK, v4.1.8.0) Best Practices workflow to remove duplicates and recalibrate base quality scores. SNVs and indels were detected using Mutect2^73^ (4.1.4.1), MuSE^74^ (1.0rc), VarDict^75^ (1.8.2), and Strelka2^76^ (2.9.2) using matched normal samples as controls. Mutect2 was used with a panel of normals composed of the complete set of matched normal. Each caller was run independently on each tumor-normal pair, and the calls were integrated using SomaticCombiner^77^. Only those SNVs detected by Mutect2 and supported by at least one of the other callers were considered for downstream analysis. In order to increase the accuracy of indel detection, only indels detected by at least two algorithms were considered for further analysis.

#### CCLE and GDSC cell line data

The mutation calls and gene expression matrix of CCLE^5^ cell lines were downloaded from DepMap data portal (https://depmap.org/portal/download/). GDSC^42^ drug sensitivity data was downloaded from the project portal (https://www.cancerrxgene.org/). To reduce the germline SNP contamination, SBS and indels were filtered with a very strict MAF cutoff of 0.001%.

### Sample selection criteria

*TCGA data:* Among MMRD tumors, a high prevalence of mutations from processes unrelated to MMRD can confound the stratification based on relative signature exposures (see below); because MMRD leads to hypermutations, this occurs most frequently when other processes causing hypermutations are active. In the TCGA data, leading sources of hypermutations are polymerase proofreading deficiency in POLE-exo or POLD1-exo mutant tumors, UV radiation, tobacco smoking, and APOBEC signatures. We clustered MMRD tumors based on their relative exposures (Supplementary Figure 14); among these clusters *L*, *Ma*, *Mb*, and *Others* contained a high prevalence of additional mutational processes. The tumors in these clusters were excluded from the comparison of SBS26-high versus –low and SBS54-high versus –low tumors. We excluded 11% of all MMRD samples in the TCGA data reducing sample counts from 413 to 367. The majority of the excluded samples had accompanying polymerase proofreading deficiency (8.2%), and among those, 82.3% were endometrial cancers. In the ICB cohorts, there were no tumors that had active additional hypermutational processes matching to the excluded clusters. Therefore, their removal in the TCGA data increases the similarity in the characteristics of the TCGA MMRD tumors to the ICB cohorts. Note that the ICB cohorts do not contain endometrial cancers. When TCGA samples are restricted to esophagogastric, colorectal, and kidney cancers that make up the majority of samples in ICB cohorts, only 1.2% of cases belonged to the excluded clusters, explaining the lack of such cases among the 95 samples in discovery and validation cohorts.

#### WGS data

For the discovery step of SV signatures samples with at least 50 SVs are selected (n = 1403), while in the second step where the de novo signatures are refitted to each sample the full PCAWG cohort (n = 2705) is used. No restriction on SV counts was applied in the results reported in Figure 3.

#### Cancer cell lines

The drug screens for 449 drugs were obtained from the GDSC project. Out of 449, 421 were tested on more than 10 MMRD cell lines and the 412 that have more than five cell lines in both the SBS26 and SBS54-high categories were used in analysis. Using the Z-scores obtained from GDSC, we performed a second Z-score transformation by setting the baseline to the average in MMRP tumors in each tissue: 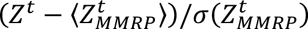, where t stands for a cancer-type of origin. Of the 1697 cell lines with WES data obtained from CCLE, 980 had drug response data. Out of 980, 81 was identified as MMRD cell lines (3 with POLE-exo mutations) and 899 as MMRP. In addition, cell lines from hematological lineages were removed as the activation induced cytidine deaminase (AID) signature^78^ frequently observed in these cell lines is very similar to the defective mismatch repair signature SBS6 and might confound the MMRD classification. Although not all cell lines were used in the analysis, we provide the MMRD status for 1697 cell lines in Supplementary Table 14 as a resource.

#### Validation cohort

In Bortolomeazzi *et al.* cohort, two samples (UH2, UH6) did not demonstrate signatures of MMRD. These samples were also not hypermutator and had low MSISensor scores (1.3, 2.5, respectively). As a result, we conclude they did not have MMRD and they were excluded from analysis. In Kwon *et al.* cohort, we only used the primary tumors and excluded any post-treatment samples.

### MMRD identification

We developed an MMRD classification method which combines MSI, quantified using either MSISensor score or indel counts at microsatellite regions, with MMRD-related SBS signatures for improved detection. MMRD score is predicted with gradient boosting classifier. Classifiers were trained on simulated panels, which are generated by downsampling TCGA data to MSK-IMPACT panel coverage and for the numbers of microsatellite loci that are covered with this panel (See Additional Methods for more detailed description). The classifier and the signatures calculated in this study (see below) are integrated in our SigMA algorithm (2.0)^27^. See Additional Methods and Supplementary Figure 1 for a detailed description and results of the validation of our MMRD classifier.

### Mutational signature analysis

#### SBS signatures

The SBS signatures (Supplementary Figure 15) were discovered using non-negative matrix factorization implemented in the SigProfiler MATLAB package (2.5.1.9)^28^, using WES (n = 413, TCGA) and WGS (n = 40, PCAWG and ICGC) MMRD and POLE-exo mutant tumors. The new signatures were matched to the COSMIC catalog (v3.2), and they were replaced with the catalog equivalent if matching was achieved. Note that in our *de novo* discovery, SBS6 distribution (Supplementary Figure 15) differed from SBS6 in the catalog (cos = 0.51). We found that this was a result of the normalization we performed on the input matrix for the NMF calculation, where we divide the mutational spectrum of each sample with the total count of SBSs. When this normalization step is not performed, SBS6 similar to that in COSMIC catalog v3.2 was discovered. Upon inspection of the per-sample exposures, this composite signature was observed to be driven by a single sample. Therefore, we believe COSMIC v3.2 SBS6 is a composite signature where we observe a mixture of SBS1 and true SBS6. Although in the COSMIC signature catalog (v3.2), SBS54 is annotated as germline contamination, we discovered it as a distinct signature in our NMF analysis on the somatic mutations from the combined WES and WGS dataset. We analyzed the VAF distribution mutations associated with SBS54 to confirm their somatic origin (Supplementary Figure 16). See Additional Methods for more information about *de novo* discovery.

MMRD signatures discovered, as described above, were then integrated into the SigMA algorithm, which is used in the refitting of signatures to calculate per-sample exposures with non-negative least squares (NNLS) algorithm (See Additional Methods). In the refitting, the MMRD SBS signatures were combined with the SBS signatures found in MMRP tumors in each tumor type (a functionality provided by SigMA).

#### Defining high signature exposures

Following the above description, the signature exposures were calculated for 175 MMRD samples (including additional MMRD samples from patients who were not treated with ICB but those that were used in the comparison of our MMRD classifier to IHC^31–33)^. The signature quantiles were defined for each signature using relative exposures, defined as the absolute exposure divided by the total number of SBSs. A sample is denoted to have a high exposure for a signature if the relative exposure is greater than the third quartile, Q3, selecting the top 75-100% of all samples. Note that potentially this threshold can further be optimized by analyzing more ICB-treated MMRD samples in the future. Here to avoid overfitting and because signature exposures do not demonstrate a clear bimodal feature we used the Q3 values as a working point. For the analysis of TCGA data, the same thresholds were used because the mutational spectra in the TCGA data to that in down-sampled panels are highly similar (Supplementary Figure 17). For WGS data, the SBS26 quartile was recalculated due to the differences in the trinucleotide frequencies. Thresholds are summarized in Supplementary Table 15.

In two of the ICB validation cohorts (Bortolomeazzi *et al*. and Kwon *et al.)*, for patients with multiple samples from pre-treatment stage, we calculated the relative exposures and whether they belong to high or low groups for each sample. The high *vs* low categories obtained from individual samples were identical for all the samples with multiple samples. In the results, each patient is only accounted for once.

#### SV signatures

The SV spectrum for each sample was calculated by counting the four types of SVs (deletion, duplication, inversion, translocation) in various segment length intervals; translocations were counted without considering segment length. The clustered SVs counts are calculated separately from the non-clustered ones. Clustered SVs were defined as false discovery rate (FDR) < 0.1, obtaining the FDR values from our previous publication^79^. The SV spectra of all PCAWG samples with at least 50 SVs were used in the de novo discovery. The exposures of discovered signatures were estimated by NNLS for all PCAWG dataset without a minimum SV restriction. SV signature exposures can be found in Supplementary Table 16.

#### Tumor type-specific correction for SV signature exposures

To account for tumor type-specific differences, we determined the mean relative exposure of SV signatures in MMRP tumors for each tumor type. Then, we subtracted the tumor type averages from the relative exposure of SV signatures in MMRD samples. Figure 3b shows the averages across samples after this subtraction, reflecting how they differ from MMRP samples. The same procedure is followed for *BRCA2*-/-samples in Figure 3c. Instead, for SVs that overlap with delayed replication fragile sites because we are comparing them to all SVs in the same tumors, the subtraction is done at a per sample level rather than tumor type averages. As a crosscheck: (1) We compared MMRD tumors in different signature groups with MMRP tumors without the correction (Supplementary Figure 8c). (2) Instead of subtracting the tumor type averages, we randomly selected two MMRP tumors of the same tumor type for each sample and compared the MMRD tumors to this randomly sampled MMRP control set. In addition to subtracting out tumor type averages, this analysis also captures the fluctuation of relative SV signature exposures in MMRP samples in addition to their average, confirming our finding (Supplementary Figure 8d,e).

### MSISensor scores and survival analysis

MSISensor scores for the discovery cohort (Samstein *et al.* ^30^) were obtained from the previous publications^32–34,36^. For the Bortolomeazzi *et al.* and Kwon *et al.* datasets in the validation cohort, MSISensor scores were calculated using MSISensor-pro (1.2.0) (https://github.com/xjtu-omics/msisensor-pro) from tumor and matched normal WES samples. These calculations were used in the combined analysis of discovery and validation cohorts (Supplementary Figure 13, and 18). For patients with multiple pre-treatment samples, we calculated the MSISensor score and we then averaged them. We found that our calculations differed from MSISensor scores reported in the other two cohorts (Samstein *et al.,* and Rousseau, Foote *et al*.) calculated from targeted gene panels, likely due to the lower total read depth in WES datasets. To combine these cohorts in downstream analysis, we shifted the MSISensor scores in the WES cohorts by adding 18 to WES scores which results in distributions that overlap with panel data (Supplementary Figure 19).

#### Selecting high vs low samples

The median, lowest (Q1), and highest (Q3) quartiles of MSISensor scores were calculated in the discovery dataset (n = 36). We compared samples with MSISensor scores larger and smaller than each threshold, referred to as MSISensor-high and low groups, respectively (Figure 1; Supplementary Figure 3) – not to be confused by MSI-low, which is used in literature interchangeably with MMRP. The thresholds used here are defined among MMRD tumors; therefore, low or high is relative to other MMRD samples.

#### Survival analyses

In the discovery cohort, none of the comparisons yielded a significant difference in OS and PFS, but the OS was somewhat shorter in samples with MSISensor score < Q1. Next, discovery and validation cohorts were combined, and the analyses were repeated, yielding a similar result (non-significant but shorter survival in samples with MSISensor < Q1; Supplementary Figure 18). However, in the combined analysis of discovery and validation cohorts, using the same threshold as in a previous publication by Mandal *et al.*^13^, a significantly shorter OS (*p* = 0.016; Supplementary Figure 20) was observed for MSISensor-low samples (MSISensor score < 21.5 < Q1). PFS and response rates were also lower for this selection, but *p*-values were not significant.

To demonstrate that MSISensor score and signature-based stratifications are independent, we built a Cox-regression model using MSISensor-low (< Q1) and SBS26 or SBS54-high status as binary covariates. We found that both selection criteria acquire statistically significant and larger than one hazard ratios of death (Supplementary Figure 13). Repeating the same analysis for PFS, the hazard ratio for SBS26 or SBS54-high selection was significant but not for MSISensor-low selection. Finally, patients that are SBS26 or SBS54-low and MSISensor-high have higher response rates compared to remaining patients (Supplementary Figure 13).

### Tumor mutational burden

TMB values were obtained from corresponding publications except for Kwon *et al.,* for which we calculated TMB (the nonsynonymous mutation counts per Mb) using our mutation calls. For patients with multiple pre-treatment samples in Kwon *et al.* and Bortolomeazzi *et a*l. cohorts, TMB average per sample is used.

#### Selecting high vs low samples

The median and quartiles for TMB were calculated in the discovery cohort (n= 55); these values were used to stratify TMB-high and low samples. TMB-high *vs* low and MSISensor-high *vs* low selections often coincide (*p* < 0.01, Fisher’s exact test). MSISensor score and TMB reflect the intensity of mutagenesis in MMRD tumors, while signature-based selection using relative exposures does not overlap (*p* > 0.05, Fisher’s exact test) with these categories and is complementary.

#### Survival analysis

We did not observe significant differences in overall and progression-free survival, and response rates among TMB-high and –low groups (defined according to Q1, median or Q3) in the discovery cohort (Supplementary Figure 3) and also in the combined analysis of discovery and validation cohorts (Supplementary Figure 18). However, when we use the same definition of TMB-low tumors as in Kwon *et al.* (TMB < 26.5 per Mb < Q1), a significantly shorter OS was observed (Supplementary Figure 20).

### Pathway analysis

#### Expression

For each sample, gene set enrichment (GSE) scores were calculated using the R package GSVA (1.44.2)^80^ on the log2-transformed expression values after filtering genes with low standard deviation and low median expression levels. Reactome^42^ and Hallmark^43^ pathways, downloaded from the MSigDB database (https://www.gsea-msigdb.org/gsea/msigdb), were analyzed. Using per sample enrichment scores, we compared samples with high *vs* low SBS26 and SBS54 exposures using the Student’s t-test. The calculation was repeated separately in each of the three major tumor types with high MMRD prevalence (COAD, STAD, UCEC) (Supplementary Table 8) to account for the tumor type-specific differences. The results presented in Figure 2 and Supplementary Figure 9 were combined by averaging the t-values in the three tumor types (Supplementary Table 7). A comparison between the pan-cancer t-values and the mean t-values is shown in Supplementary Figure 21. For the pan-cancer t-test results, two approaches for multiple hypothesis testing were followed using (1) simulations with random sampling and (2) Benjamini-Hochberg procedure (Additional Methods; Supplementary Table 7; Supplementary Figure 22).

#### Mutations

We calculated the difference in the frequency of alterations (truncating and nonsynonymous mutations, copy number amplification, and deletions) for all genes and used the differences to infer the gene set enrichment scores for the Reactome pathways. The fraction of samples with mutations in the high *vs* low groups for SBS26 and SBS54 were calculated for each gene. The differences in the fraction of samples with mutation (fraction in samples with high exposure minus fraction in samples with low exposure) were used to calculate the GSE scores with the gsePathway function from the R package ReactomePA (1.40.0)^81^. You can find the code sniplets used for the above calculation can be found in the Additional Methods document.

### Prediction of neoantigen counts

For each SBS and indel, translations over all three possible frames were simulated from sequences consisting of the mutation along with the 36 bases flanking each side. Translations were truncated at premature stop codons and broken up into all remaining 8-12 amino acid peptides. Peptides from the unaffected sequences were similarly isolated as a control set. For each sample, nanomolar binding affinities between the sample’s MHC alleles and the peptides originating from SNV and indels were calculated using netMHCpan^82^ (4.1). For mutations whose flanking window crosses a splice junction, all exon-exon junction sequences annotated in the ENSEMBL hg19 assembly were considered. For SBSs, each mutant peptide’s affinity was compared to the affinity of the corresponding wild-type amino acid sequence. For example, given an SBS, e.g. A>T, the wild-type peptide arising from any genomic sequence containing the A was compared to the peptide arising from the corresponding genomic coordinates with the genomic sequence altered by the T. In the case of indels, the affinities of any peptide generated by completely new genomic sequences or reading frames were compared as a population to the population of the affinities of peptides generated from the wild-type sequence. Strong neoantigens are those within the top 0.5% of all peptides. We simulated neoantigens from all possible isoforms whose coding or splicing sequence might be altered. For example, if an indel affects an exon found in multiple isoforms, we simulated all frameshifted sequences from isoforms involving the indel.

### Determining samples with HLA LOH

We performed HLA typing for tumor and matched normal samples in TCGA data using Polysolver (https://software.broadinstitute.org/cancer/cga/polysolver_download). We determined HLA-LOH by comparing the sequence of most likely HLA alleles estimated by Polysolver^83^ in tumors to that in the normal samples. If the sequence differences in the tumor were more than 5 times smaller than the distance to the closest normal allele and if this distance was < 250, we considered the HLA to have LOH. We compared the GSE score for IFN-γ (Hallmark) in samples with a loss of all three HLA genes versus those only a subset has been lost. We found that sampeles with LOH of all three HLA genes had lower GSE scores for IFN-γ pathway (Hallmark), while those with a loss of only a subset of the genes had a higher IFN-γ expression, and did not differ significantly from samples with no detected LOH. Therefore, we only considered the loss of all three genes as HLA-LOH in the downstream analysis.

### Identifying subclonal mutations

If purity corrected VAF of a mutation is in agreement with the
 expected VAF based on the local copy number state, it was tagged as clonal. Using binomial distributions, we tagged mutations with less than 5% chance of being to a clonal as subclonal candidates. We revised the classification of mutations that were in the VAF boundary between subclonal and clonal mutations. We calculated the difference for *VAF* · *CN*_total_ values of each mutation to the average of the same measure in clonal (considering only those on one copy of the chromosomes), and subclonal groups and we divided by the respective standard deviation. If a subclonal mutation was closer to clonal group, and if likelihood of being clonal was > 1%, we recategorized it as a clonal mutation. The remaining subclonal mutations were clustered to determine individual subclones using hierarchical clustering. The average of *VAF* · *CN*_*total*_ values for mutations in each subclonal cluster were then used to estimate the cancer cell fraction of subclones.

### Immune cell proportions

We used CIBERSORTx^84^ in order to deconvolute tumor microenvironment (TME) cell-type populations from the TCGA bulk RNA sequencing data. As a reference for the cell populations to deconvolute, we built a TME signature matrix using recently reported pan-cancer single-cell transcriptomics data^85^. We downloaded the expression count data from the Lambrechts lab website (http://blueprint.lambrechtslab.org) and merge them into a unique Seurat object (using the *Seurat* R package)^86^. We labeled the cells using the fine cell type calls kindly given by the authors upon request then subset the Seurat object in order to have a maximum of 400 cells per cell type. The matrix of count data was used as an input in CIBERSORTx to build a gene expression signature matrix (by disabling the quantile normalization parameter, other parameters set to default). We then run the deconvolution in absolute mode using the TCGA RNA sequencing count data as mixture matrix (500 permutations, disabled quantile normalization). The relative mode could be rescued by summing all populations to 1. We then compared the cell population frequencies or abundance by performing comparisons between different groups.

### Statistical tests

The Kaplan Meier overall survival curves were compared using the log rank test, and the significance of the hazard ratios was determined using the Wald statistics implemented in the R package the survival. The geneset enrichment scores and single gene expression levels were compared using double sided t-test (*t.test* function implemented in the R package core stats).

## Code availability

The code and the MMRD classifiers are available in SigMA R package at https://github.com/parklab/SigMA. The code for neoantigen predictions can are available at https://github.com/parklab/mutagenesis_tools.

## Ethics and inclusion statement

This work is conducted according to Global Code of Conduct, and does not exploit local communities.

## Data Availability

All data is obtained from previous publications and data produced as a result of their analysis are contained in the manuscript

## ACKNOWLEDGEMENTS

This work was funded by grants from the National Institutes of Health (R01CA269805 to P.J.P.), Ludwig Center at Harvard (P.J.P.), Cancer Research UK Grand Challenge and the Mark Foundation to the SPECIFICANCER team (P.J.P.), and Swedish Research Council (2020-00583 to V.L.). Authors would like to thank Luis Diaz for his assistance with the colorectal cancer ICB datasets, Deborah Schrag^6^, Michael Yevgeniy Tolstorukov^6^, Elizabeth F. Cohen^6^ for sharing the GENIE colorectal cancer data which was used to update the overall survival for two patients, and Panagiotis Konstantinopoulos for the insightful discussions.

