## Supplementary_Figures for "Predicting response to immune checkpoint blockade therapy among mismatch repair-deficient patients using mutational signatures"

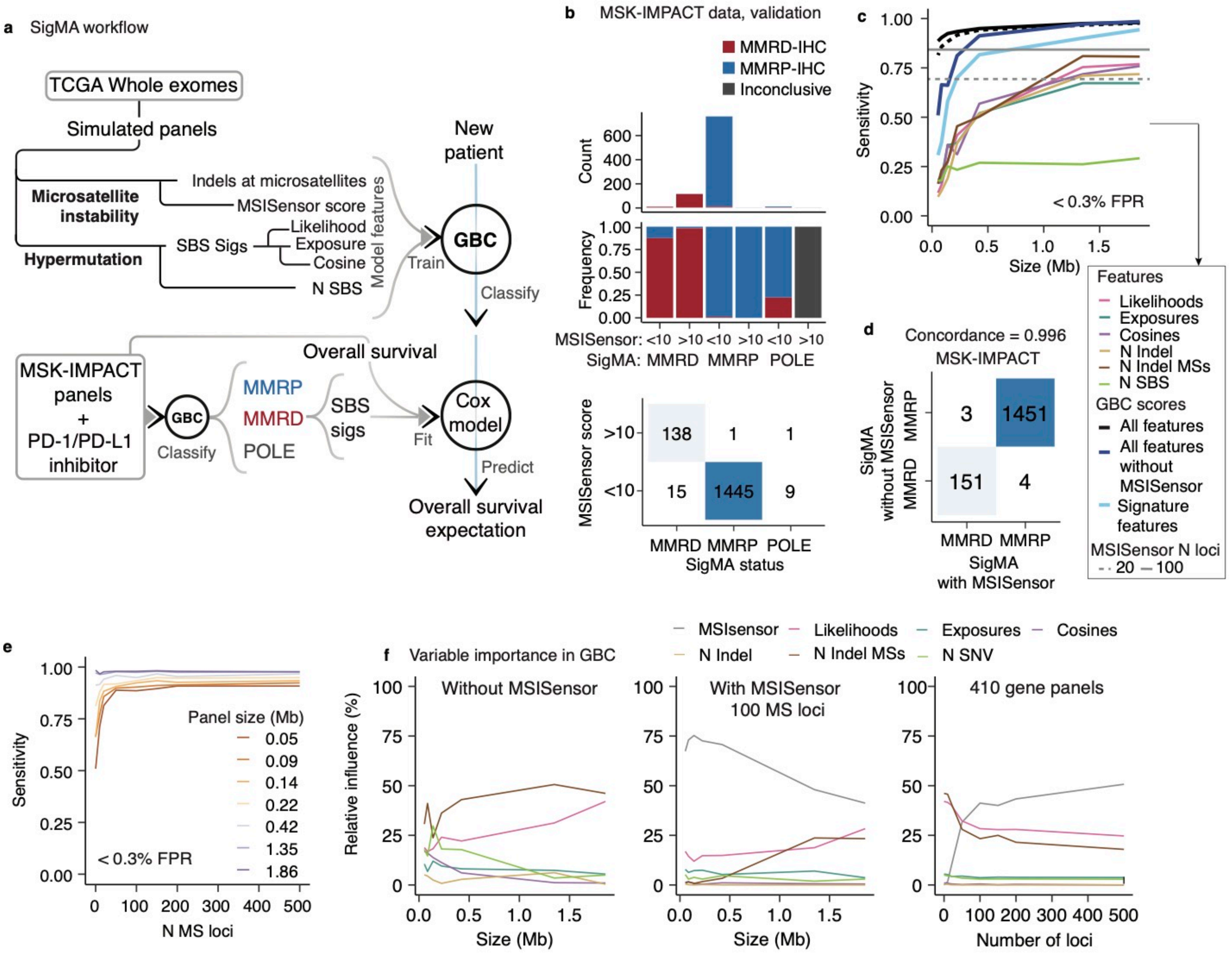


**Supplementary Figure 1. Performance of the SigMA algorithm. a.** The MMRD detection workflow implemented in the SigMA package is shown. To simulate SBSs that would be detected by gene panels, TCGA whole-exomes are down-sampled by selecting those that fall within the target regions of different panels. MSISensor scores are simulated by random sampling, given N loci and the fraction of MSs with mutations in the TCGA data. Features, including indel counts at repeat regions and signature-related measures, are calculated from the simulations. The signature-related measures are likelihoods of MMRD signature clusters (Supplementary Figure 7), cosine similarities and exposures of MMRD/POLE signatures (of SBS6, 10, 14, 15, 20, 21, 26 54), and finally, the likelihoods ratio comparing the likelihood of that specific decomposition to an alternative solution. Taking the MMRD classes in the TCGA data as the true reference, a gradient boosting classifier is trained for the simulated panels. **b.** The MMRD categories by SigMA are compared to MMR gene immunohistochemistry-based classification and MSISensor score-based selection (MSISensor score > 10, Middha *et al.*, *JCO PO*, 2017). The colors indicate the IHC status, and the three SigMA categories are further subdivided into two based on the MSISensor score. **c.** The sensitivity as a function of panel size for SigMA algorithm trained that combines signatures and indels (with and without MSISensor scores) are compared to signature-only classification and classification with each feature alone. The MSISensor score-based classification for two loci is shown as vertical lines as it is an independent simulation parameter. **d.** The confusion matrix for MMRD classification with SigMA with and without MSISensor score. **e.** Sensitivity of SigMA as a function of the number of MS loci for panels of different sizes. **f.** The relative influence of features for the GBC classifiers. (Left) as a function of panel size for training without MSISensor scores. (Middle) as a function of panel size for MSISensor loci and 100 MS loci, (Right) as a function of the number of MS loci for MSK-IMPACT panels.

**a** Basic cohort statistics

| | | All | MMRP | MMRD | T>C high | T>C low | SBS26/SBS54 high | SBS26/SBS54 low |
|---|---|---|---|---|---|---|---|---|
| Number of samples | | 1662 | 1607 | 55 | 27 | 28 | 24 | 31 |
| Drug | PD-1/PD-L1 | 1307 | 1255 | 52 | 25 | 27 | 22 | 30 |
| | CTLA-4 | 99 | 98 | 1 | 1 | 0 | 1 | 0 |
| | Combo | 256 | 254 | 2 | 1 | 1 | 1 | 1 |
| Age | <30 | 50 | 48 | 2 | 1 | 1 | 1 | 1 |
| | 31-50 | 284 | 269 | 15 | 8 | 7 | 7 | 9 |
| | 51-60 | 416 | 405 | 11 | 6 | 5 | 6 | 5 |
| | 61-70 | 499 | 481 | 18 | 10 | 9 | 8 | 10 |
| | >71 | 226 | 218 | 8 | 2 | 6 | 2 | 6 |
| Cancer type | Bladder | 214 | 207 | 7 | 2 | 5 | 1 | 6 |
| | Breast | 45 | 45 | 0 | 0 | 0 | 0 | 0 |
| | Unknown Primary | 90 | 8 | 1 | 0 | 1 | 0 | 1 |
| | Colorectal | 110 | 78 | 32 | 14 | 18 | 14 | 18 |
| | Esophagogastric | 126 | 114 | 12 | 8 | 4 | 7 | 5 |
| | Glioma | 117 | 116 | 1 | 1 | 0 | 1 | 0 |
| | Head and Neck | 138 | 137 | 1 | 1 | 0 | 0 | 1 |
| | Melanoma | 321 | 321 | 0 | 0 | 0 | 0 | 0 |
| | NSCLC | 350 | 350 | 0 | 0 | 0 | 0 | 0 |
| | Renal Cell Carc. | 151 | 150 | 1 | 1 | 0 | 1 | 0 |
| Gender | Female | 628 | 597 | 31 | 16 | 15 | 16 | 19 |
| | Male | 1034 | 1010 | 24 | 11 | 13 | 11 | 12 |

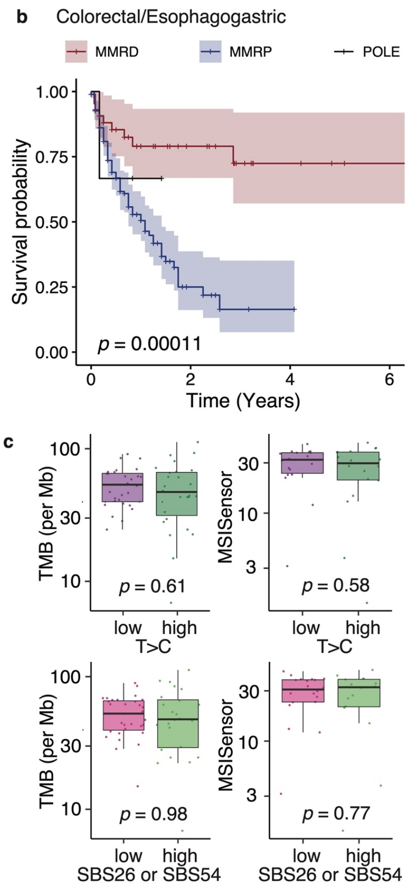


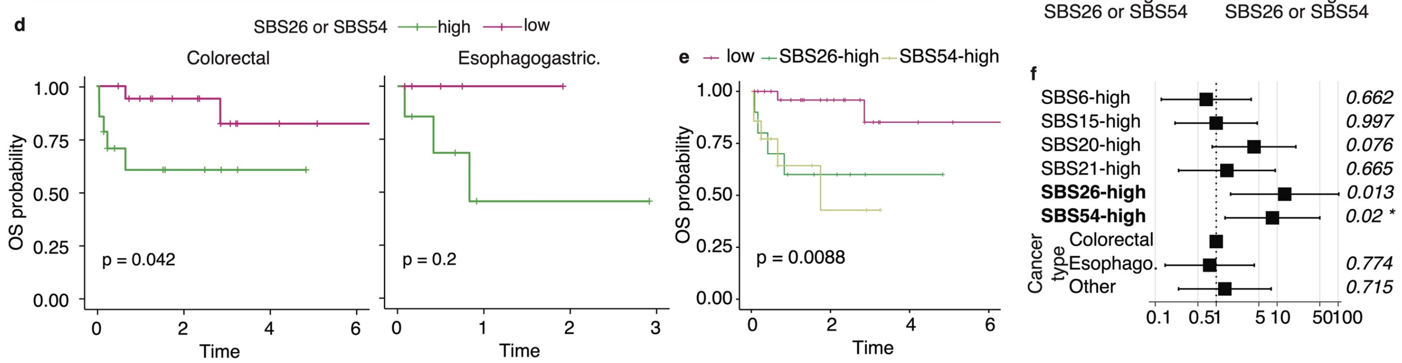


**Supplementary Figure 2. a.** Basic statistics across cohorts. **b.** Kaplan Meier (KM) curves for MMRP, MMRD, and POLE samples using only the colorectal and esophagogastric cancers. Significance estimated with log-rank test. **c.** Comparison of MSISensor score and TMB in T>C-high versus T>C-low categories and the selection using SBS26 or SBS54 (none of the distributions were significantly different, t-test). **d.** KM curves comparing SBS26-high or SBS54-high sample to the rest for patients that belong to Colorectal and Esophagogastric cancers separately in Samstein *et al.* cohort. **e.** KM curve showing SBS26 and SBS54-high groups separately in the discovery cohort (Samstein *et al.)*. **f.** Cox-regression analysis, including tumor type and age as potential confounding factors, shows that signatures still have a significant hazard ratio. All the tumors that do not belong to colorectal or esophagogastric cancers are grouped in the *other* category. For all the panels, OS information obtained from Samstein *et al.* has been updated with more up-to-date OS values from Rousseau, Foote *et al.* cohort (n = 26) and the GENIE Colorectal cancer working group (n = 2). For panels a-c, f, g, and I, the p-values for KM curves are obtained with log-rank tests for the KM figures and likelihood-ratio-tests for the hazard ratios. The shaded areas in panels a, f, g, and i indicate the 95% CI.

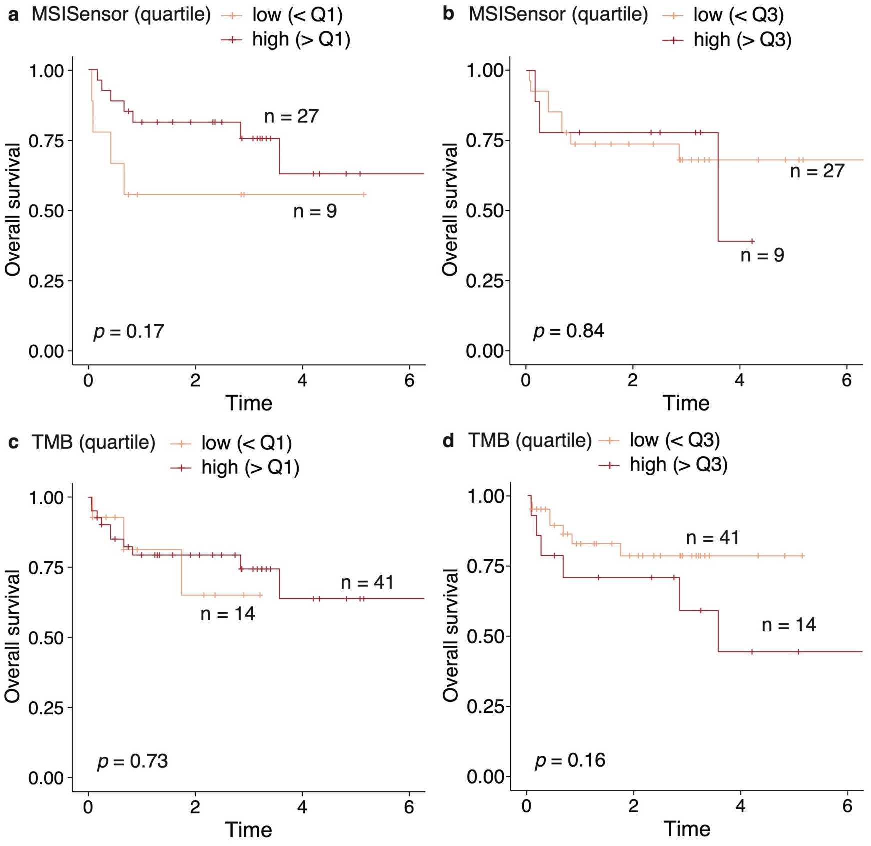


**Supplementary Figure 3. Overall survival comparison for alternative thresholds** Overall survival for stratification with **(a-b)** TMB **(c-d)** MSISensor scores. The thresholds are set at quartiles, left stratifying at the first quartile (Q1) and right stratifying at the third quartile (Q3). The black lines indicate samples with higher values of the corresponding variables. For all panels, the p-values for KM curves are obtained with log-rank tests.

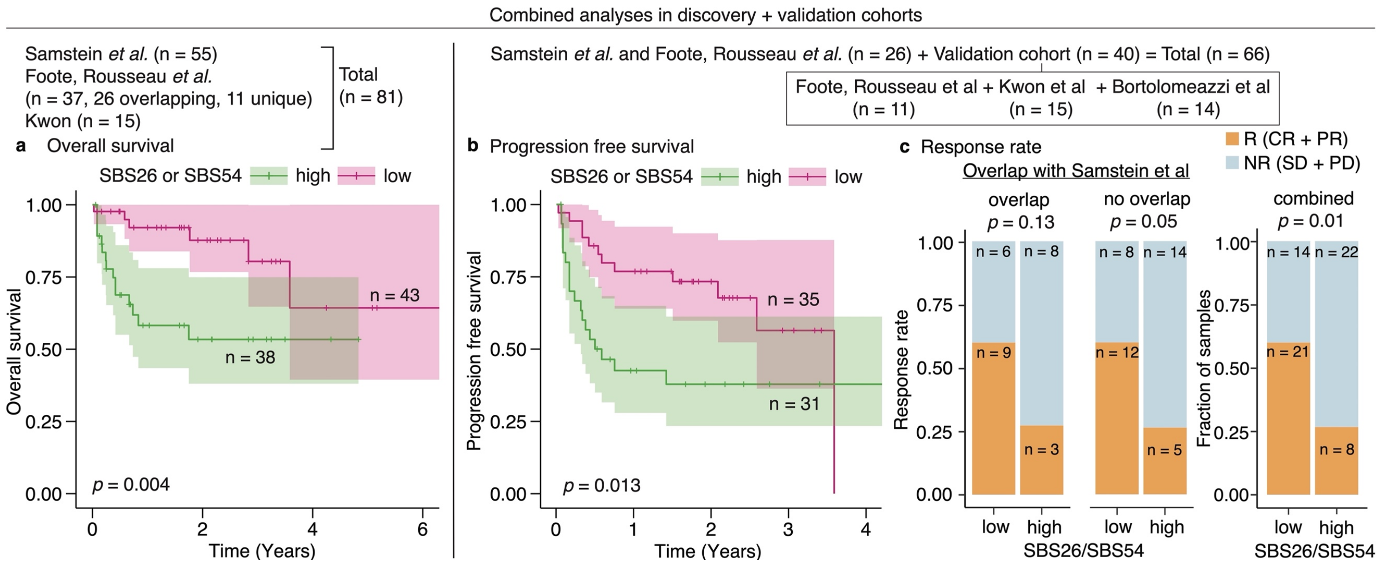


**Supplementary Figure 4. Combined analyses of validation and discovery cohorts.** Note that not all the patients in the validation cohort had OS information and not all the patients in the discovery cohort had PFS information. Number of patients from each dataset are noted on top of the panels**.** KM curves comparing SBS26-high or SBS54-high samples to low cases for overall survival **(a)** and progression free survival **(b)**. **c.** Response rate compared between the same signature-based classes: For samples that are reported by both Samstein *et al.* and Rousseau, Foote *et al.* which were not reported in Figure 1 (left), additional samples in the validation cohort which were reported in Figure 1 (middle) and both combined (right).

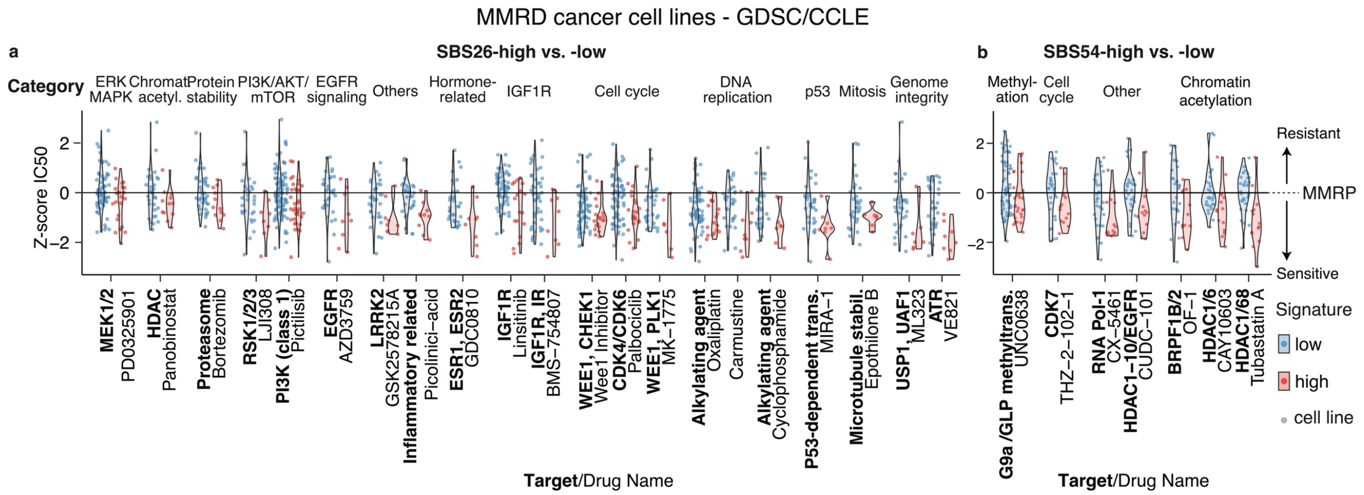


**Supplementary Figure 6. Drugs to which MMRD tumors are sensitive to depends on SBS signatures.** The Z-scores distributions for drugs with a significant sensitivity increase ($p < 0.05$, t-test, no multiple hypothesis testing correction) in the SBS26-high **(a)** and SBS54-high **(b)** categories. Drugs are ordered based on their category, and such that average t-statistics decreases by going from left to right (drugs with the largest difference between the groups are shown on the right).

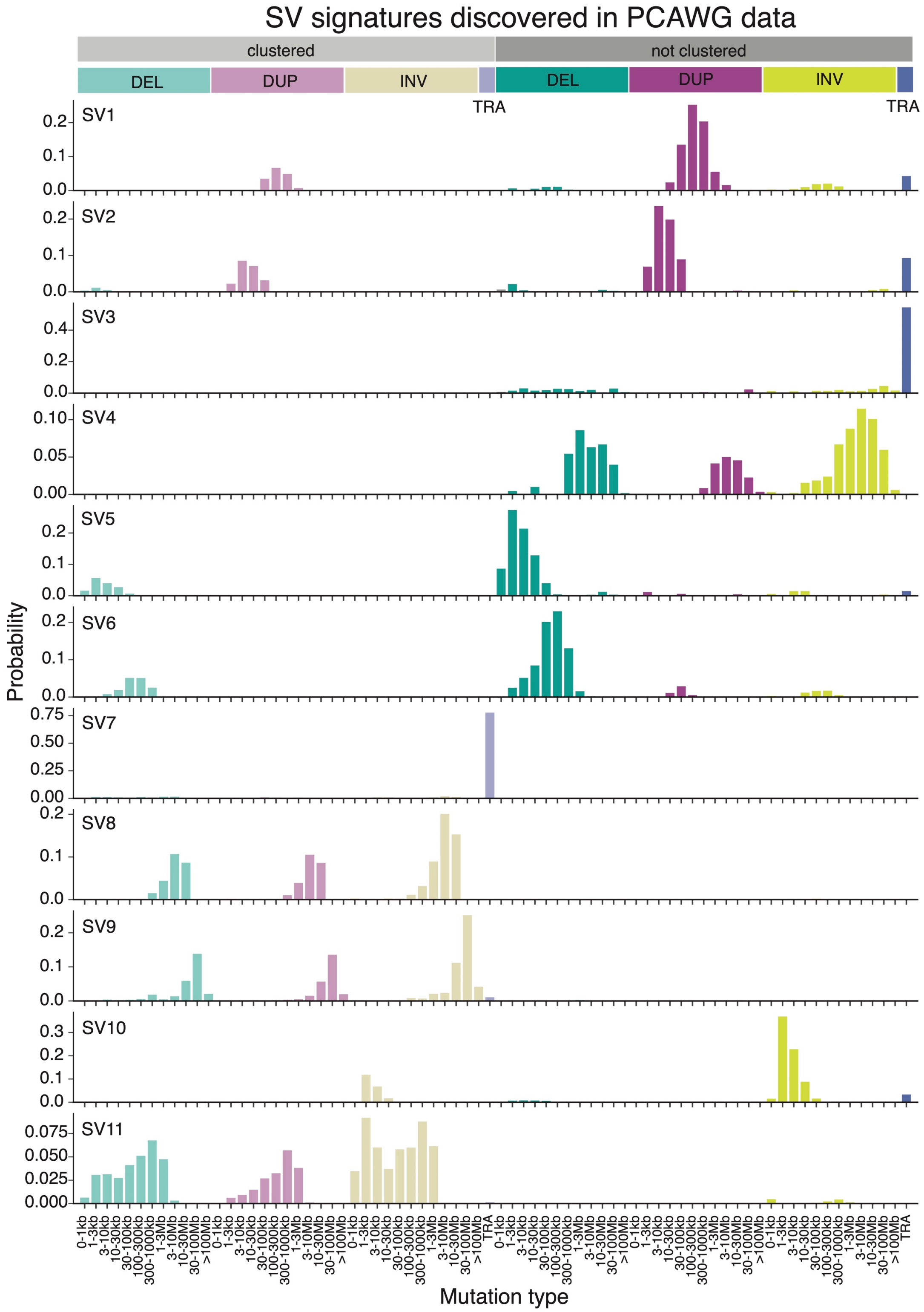


**Supplementary Figure 7. WGS MMRD analysis.** The SV signature distributions; SV types are represented by colors; clustered (left) and nonclustered (right) SVs are represented with alpha values; each SV type is split according to SV size. In Figure 3, only the second half of the distributions (nonclustered portion) are shown because the clustered SVs make little contribution.

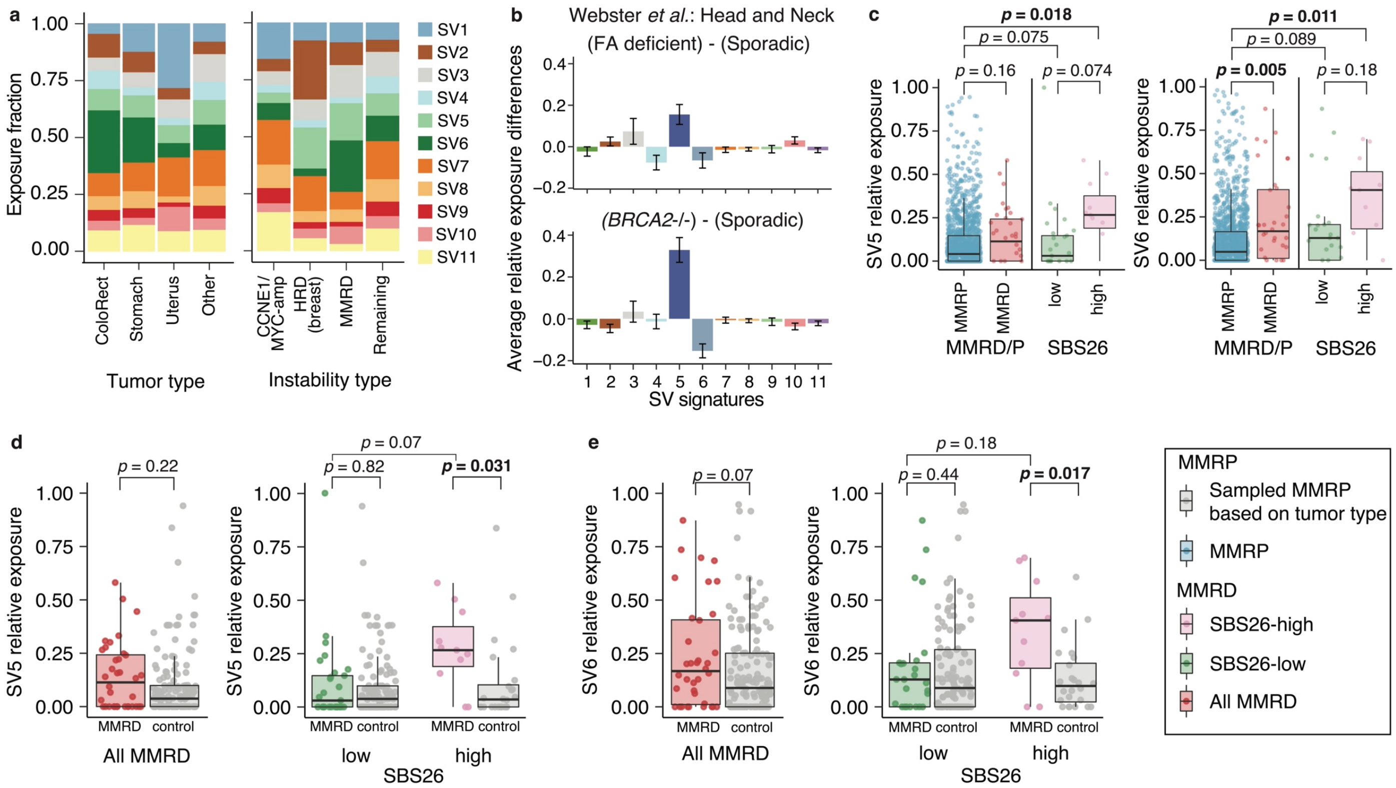


**Supplementary Figure 8. a.** The fraction of SV signatures exposures in three major tumor types associated with MMRD (COAD: ColoRect, STAD: Stomach, UCEC: Uterus) and others (Left) and comparing MMRD to breast cancer HRD samples and tumors with CCNE1/MYC amplifications across all tumor types (Right). **b.** As in Figure 3b-d, average relative exposure of SV signatures in head and neck cancers are compared between Fanconi anemia deficient and sporadic samples (top) and BRCA2-/- and sporadic samples (bottom) using data from Webster *et al. Nature* 2022. **c.** The SV5 and SV6 relative exposures are compared between MMRP and MMRD and among MMRD between SBS26-high versus low tumors; *p* values are calculated with a t-test, and significant values are emphasized with bold font. **d-e,** In order to account for the tumor type differences between MMRD and MMRP tumors as well as between SBS26-high versus –low categories, for each sample, we subsampled with replacement from MMRP tumors restricting to that tumor type is performed to generate the control group. The SV5 and SV6 relative exposures are compared between MMRD and control, SBS26 high and low MMRD to control tumors, in panels C and D, respectively.

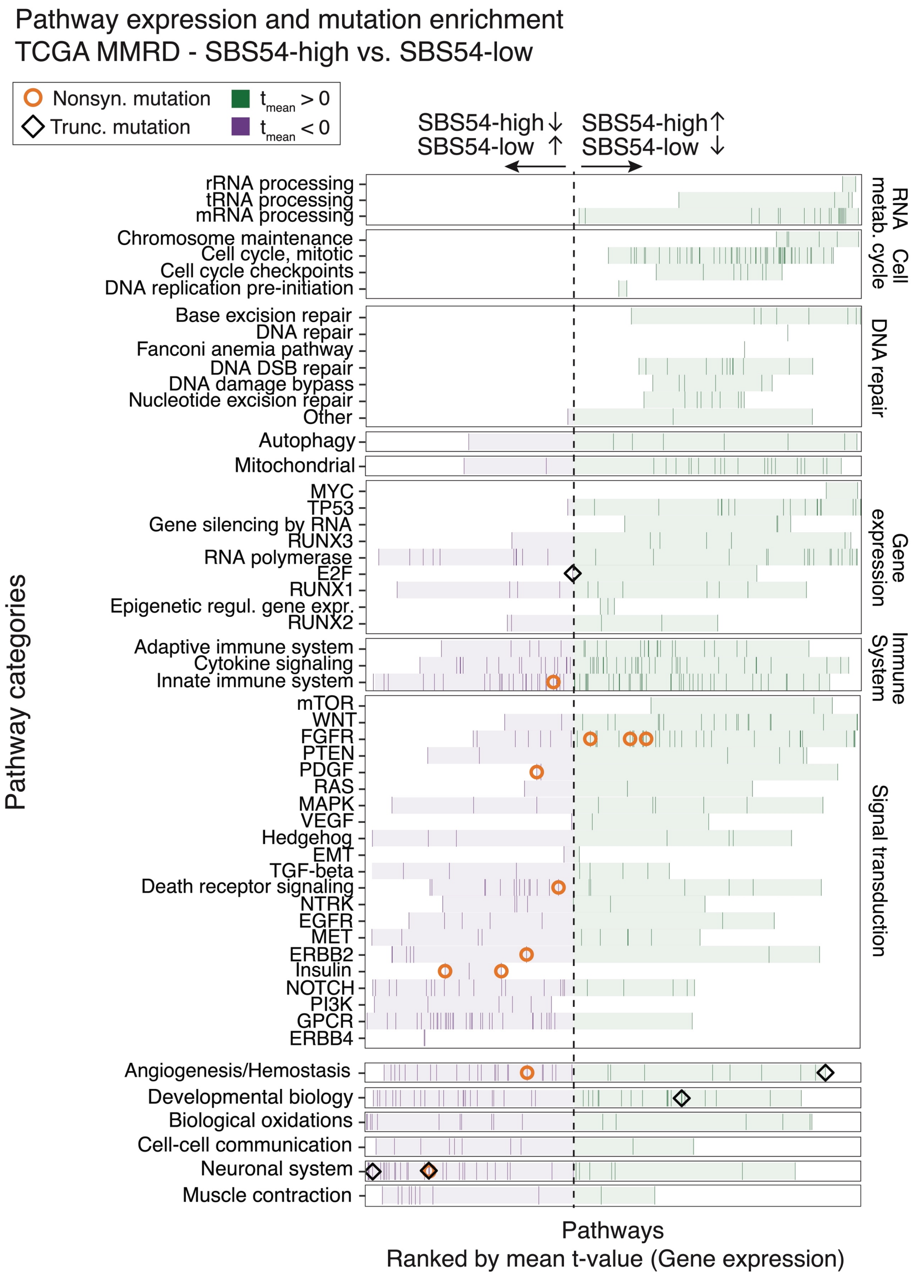


**Supplementary Figure 9. SBS54 differential pathway expression.** Same as Figure 2A but for comparison of SBS54-high and –low tumors. Pathways are ordered based on the rank of the t-value obtained from the comparison of gene set enrichment scores (Reactome[42] and Hallmark[43] gene sets obtained from MSigDB) between SBS54-high and low TCGA samples. All pathways are shown, and the rank of each pathway is denoted as a vertical line (purple or green based on whether the t-value is > 0 or < 0). Pathways are grouped in each row according to their categories (larger font blue) and subcategories (smaller font black). The shading covers the range defined by the lowest and highest ranks in that (sub)category. Significant enrichments ($p < 0.05$) in mutation frequencies in pathways in SBS26-high tumors are also marked for different types of alterations (circle: nonsynonymous mutation, diamond: truncating mutation). No significant CN-loss enrichment was observed.

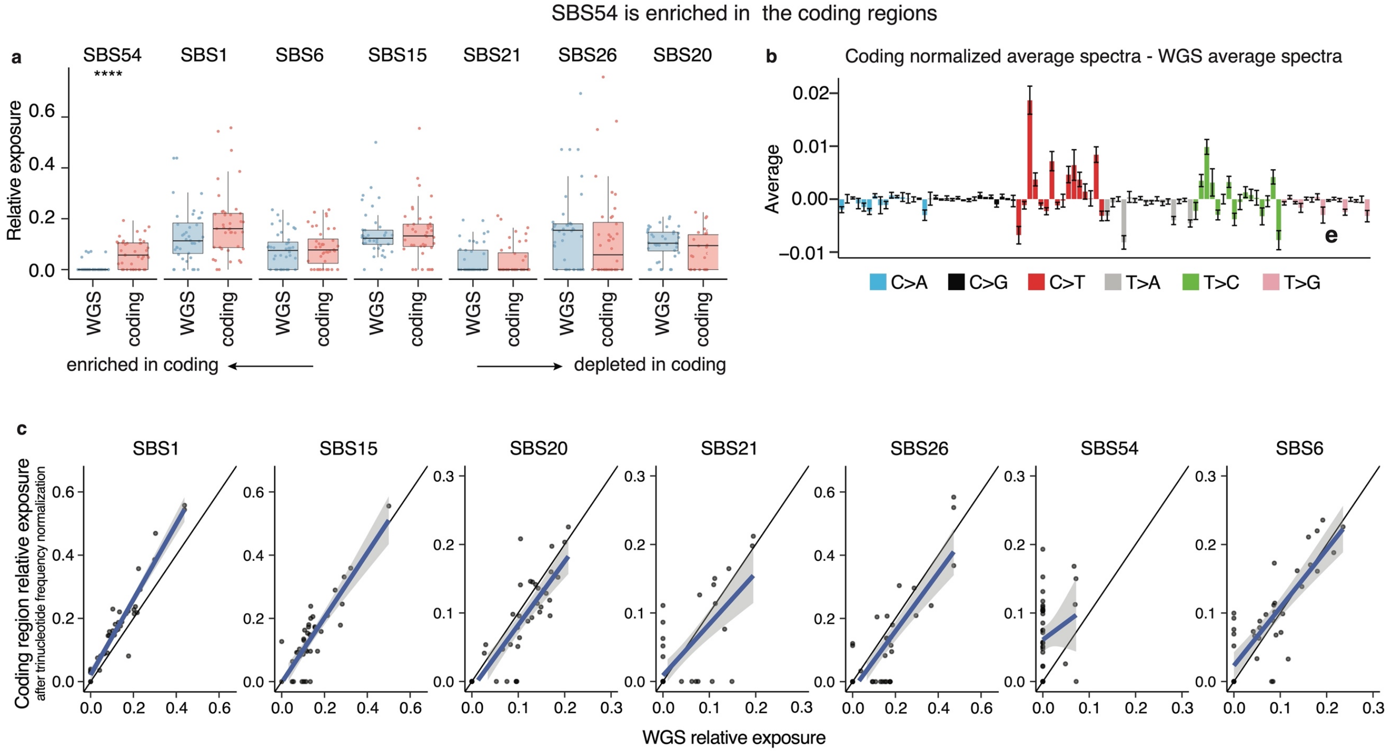


**Supplementary Figure 10. SBS54 in enriched in coding regions a.** Comparison of SBS54 exposure between coding region and whole genome; ****: $p$ < 0.0001 t-test. **b.** The difference between coding region and WGS mutational spectra is averaged across samples. The error bars indicate the standard error. **c.** Scatter plot of SBS54 exposure in coding region and whole genome. The solid black line is the diagonal showing that SBS54 distribution deviates from it. The blue line is the linear regression curve and black shaded areas are the 95% CIs. In all panels, the coding spectrum is reweighted for the differences in trinucleotide frequencies.

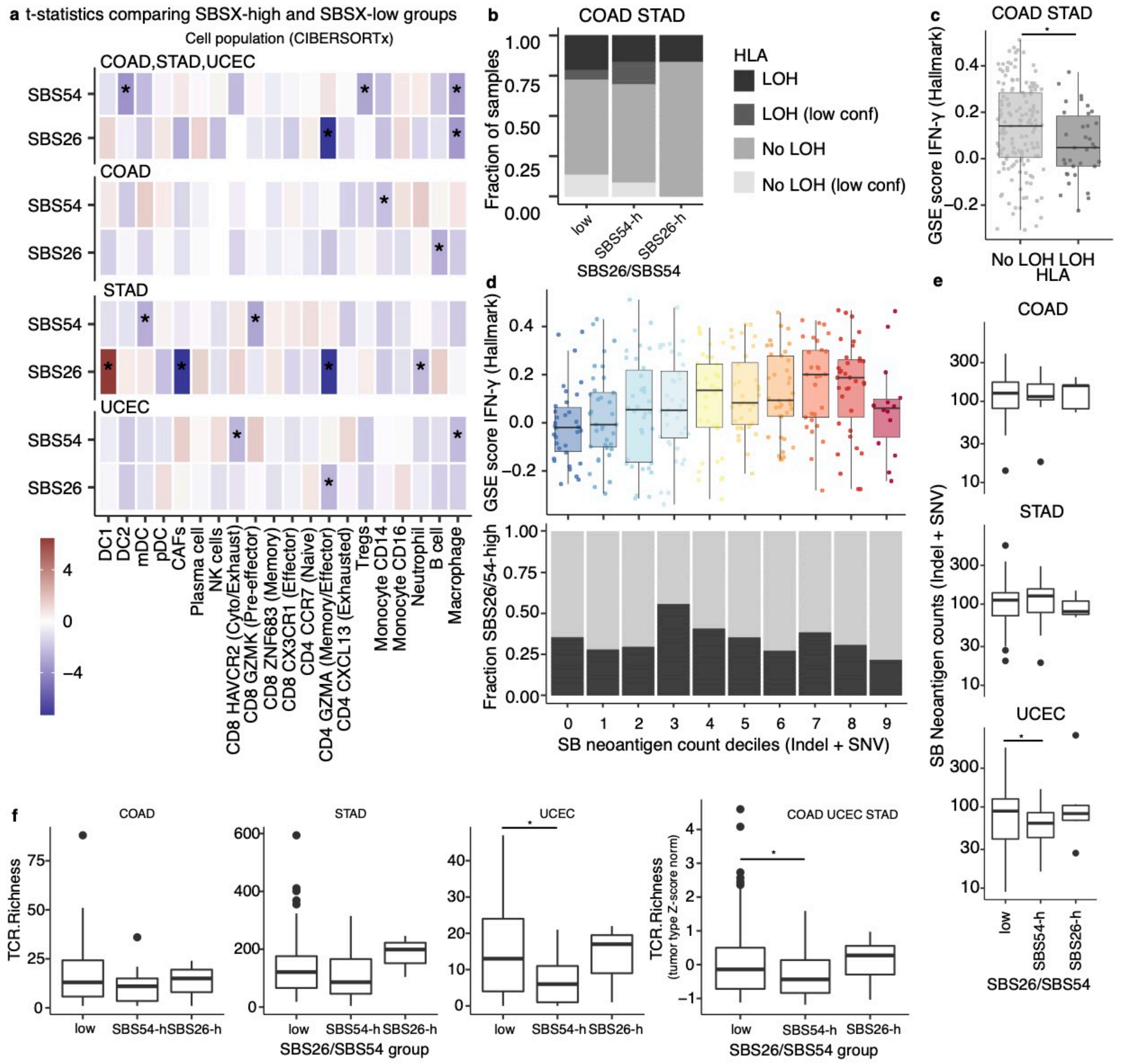


**Supplementary Figure 11. Comparison of immune landscape of SBS26/SBS54-high and low tumors. a.** Immune cell proportions estimated by CIBERSORTx from RNA-seq data are compared between SBS26-high/low and SBS54-high/low groups using t-test. For the comparison combining COAD, STAD and UCEC, first the values are normalized by a Z-score transformation using mean and standard deviation in each tumor type. The fill color demonstrates t-statistics, where negative values indicate a down regulation in the high compared to low group. **b.** Fraction of samples with LOH on HLA genes in COAD and STAD tumor types, LOH indicates loss of all three (A, B, C) HLA genes. **c.** The gene set enrichment (GSE) score calculated by GSVA package for Hallmark IFN- γ geneset obtained from MSigDB is compared between samples with HLA LOH and those without. Low confidence cases are excluded in this comparison. **d.** Top: The the IFN- GSE scores are shown in deciles of strong binding (SB) neoantigen counts, demonstrating a moderate increase with increasing number of neontigens. Bottom: The fraction of samples that are either SBS26 or SBS54-high in the neoantigen decile bins. **e.** SB neoantigen counts compared between SBS26/54 high and low cases. **f.** The TCR Richness metric obtained from Thorsson *et al.* is compared between SBS26/54 high and low groups. There is a large tumor type dependence in this metric and while combining the three major tumor types into a single bin a Z-score normalization using mean and SD in each tumor type is applied. In all the panels * indicates $p < 0.05$ for t-test.

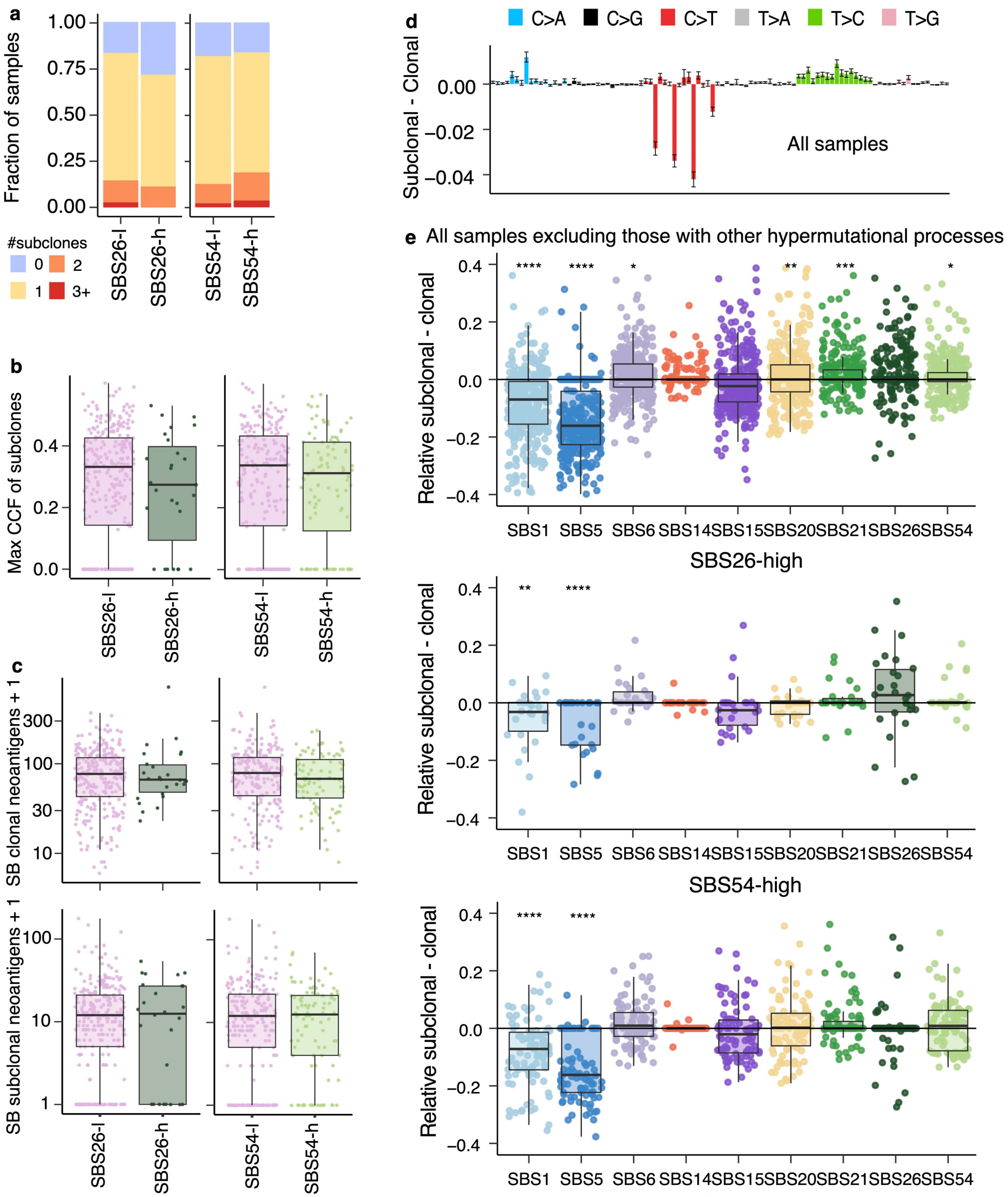


**Supplementary Figure 12. Clonality of signatures. a.** Fraction of samples with subclones, where fill color indicates number of subclones. **b.** Cancer cell fraction (CCF) (Left) and SNV count (Right) ratios between subclonal and clonal mutations in SBS26/54 high and low groups are compared. No significant difference was observed **c.** Clonal and subclonal strong binding (SB) neoantigen counts. **d.** Average difference between the normalized subclonal and clonal mutational spectra in all tumors, and SBS26-high and SBS54-high tumors. The error bars show the standard error. None of the comparisons are significant. **e.** Only samples with total Subclonal SBS/Clonal SBS counts > 0.05 are included. Difference between subclonal and clonal relative exposures in all, SBS26-high and SBS54-high samples, respectively, from top to bottom. In panels c, d $p < 0.05$: *, $p < 0.01$: **, $p < 0.001$: ***, $p < 0.0001$: *** calculated with t-test.

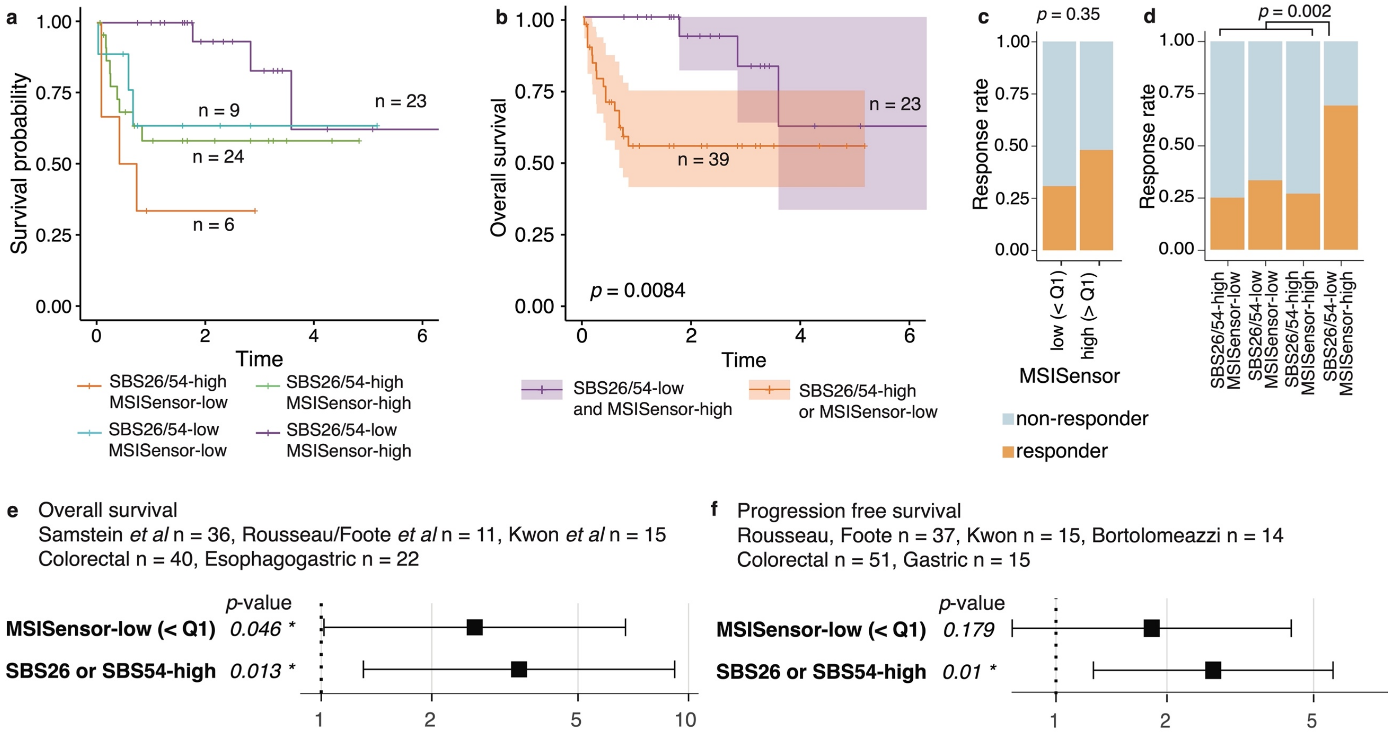


**Supplementary Figure 13. Combined survival analysis in discovery and validation cohorts stratified based on SBS26 or SBS54-high and MSISensor-low a.** KM curve of OS comparing four strata defined according to SBS26/54 and MSISensor score. MSISensor-low refers to samples with MSISensor score < Q1. **b.** KM curve of OS comparing SBS26/54-high and MSISensor-low tumors to others. In panels a-d all samples in discovery and validation cohorts with the available information (Samtein *et al.* n = 55, Rousseau, Foote *et al.* n = 10) were combined. **c.** Response rate in MSISensor quantiles **d.** Response rate in the same strata as in panel c; *p* value is obtained with Fisher's test. For panels a-c, the p-values for KM curves are obtained with log-rank tests, and shaded areas indicate 95% confidence intervals. The p-values for hazard ratios in Cox regression was obtained by likelihood test. **e-f.** Hazard ratios obtained by Cox regression with MSISensor and SB26/SBS54 based binary classification, and tumor type as covariates for OS **(e)** and PFS **(f)**, respectively

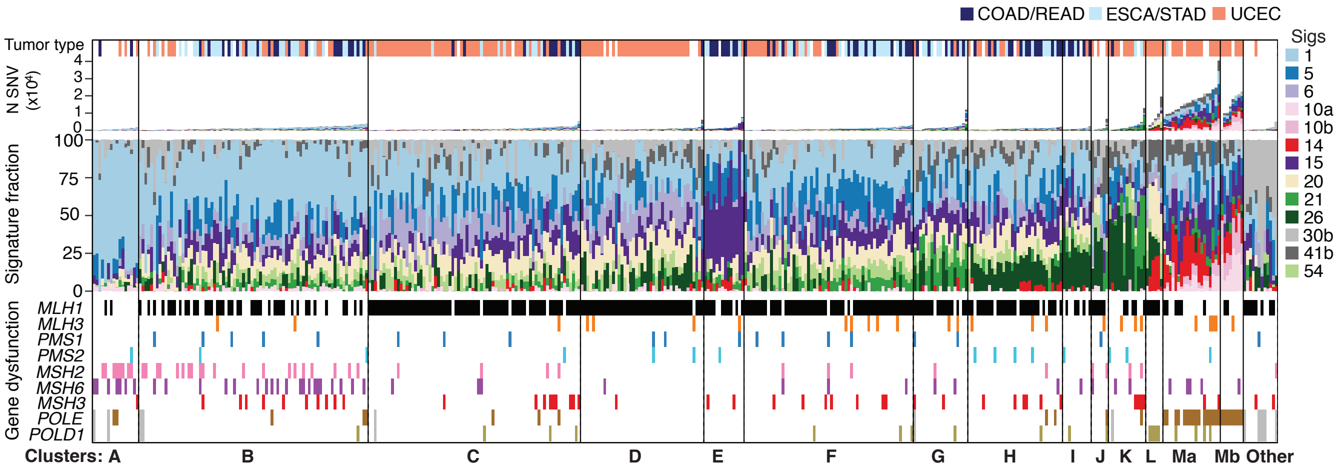


**Supplementary Figure 14. SBS signatures and MMR dysfunctions in the TCGA dataset.** Each vertical entry is a sample. Samples are first clustered according to SBS signature exposures and within each cluster sorted according to the number of SBSs. From top to bottom, (1) tumor types for the three major categories are marked, and the rest of the tumor types are shown in white. (2) Number of single base substitutions colored by the signatures according to the exposure of each signature. (3) The relative exposure of SBS signatures in each sample. (4) The MMR gene dysfunctions and mutations on exonuclease domain of POLE and POLD1 samples are annotated. (5) Letters identify the clusters.

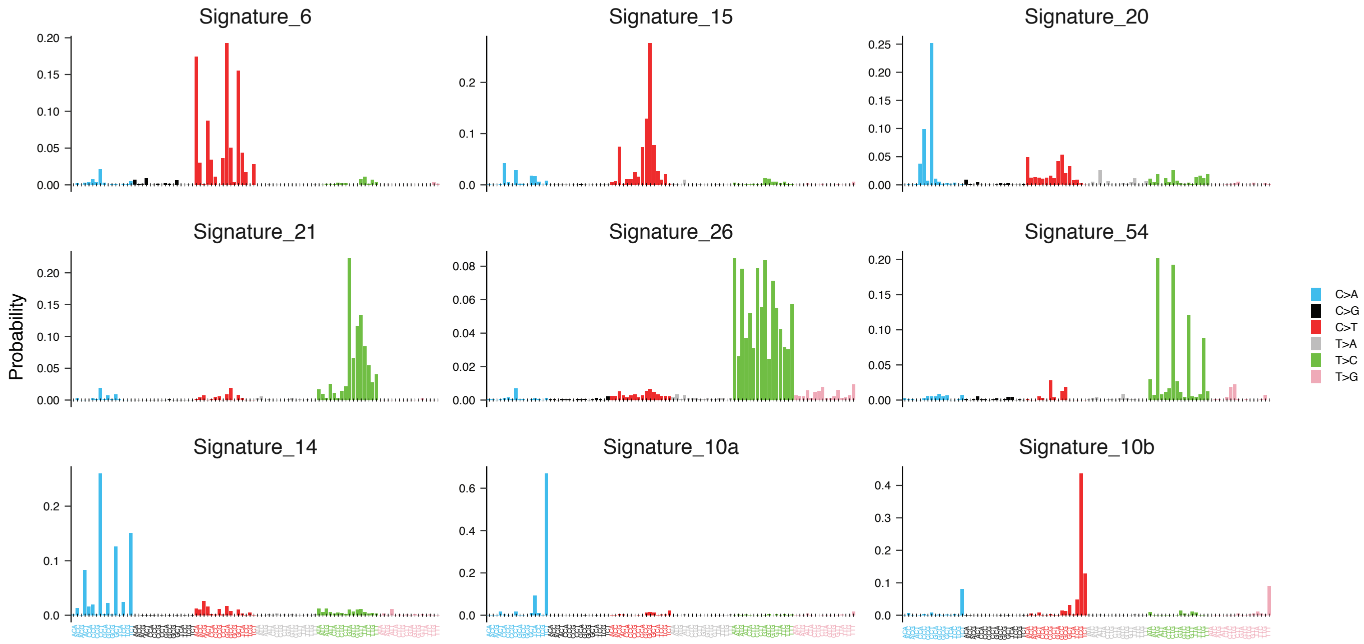


**Supplementary Figure 15. SBS signature distributions.** The MMRD-related SBS signature distributions included in the SigMA method. SBS6 has been updated with respect to the COSMIC catalog (v3).

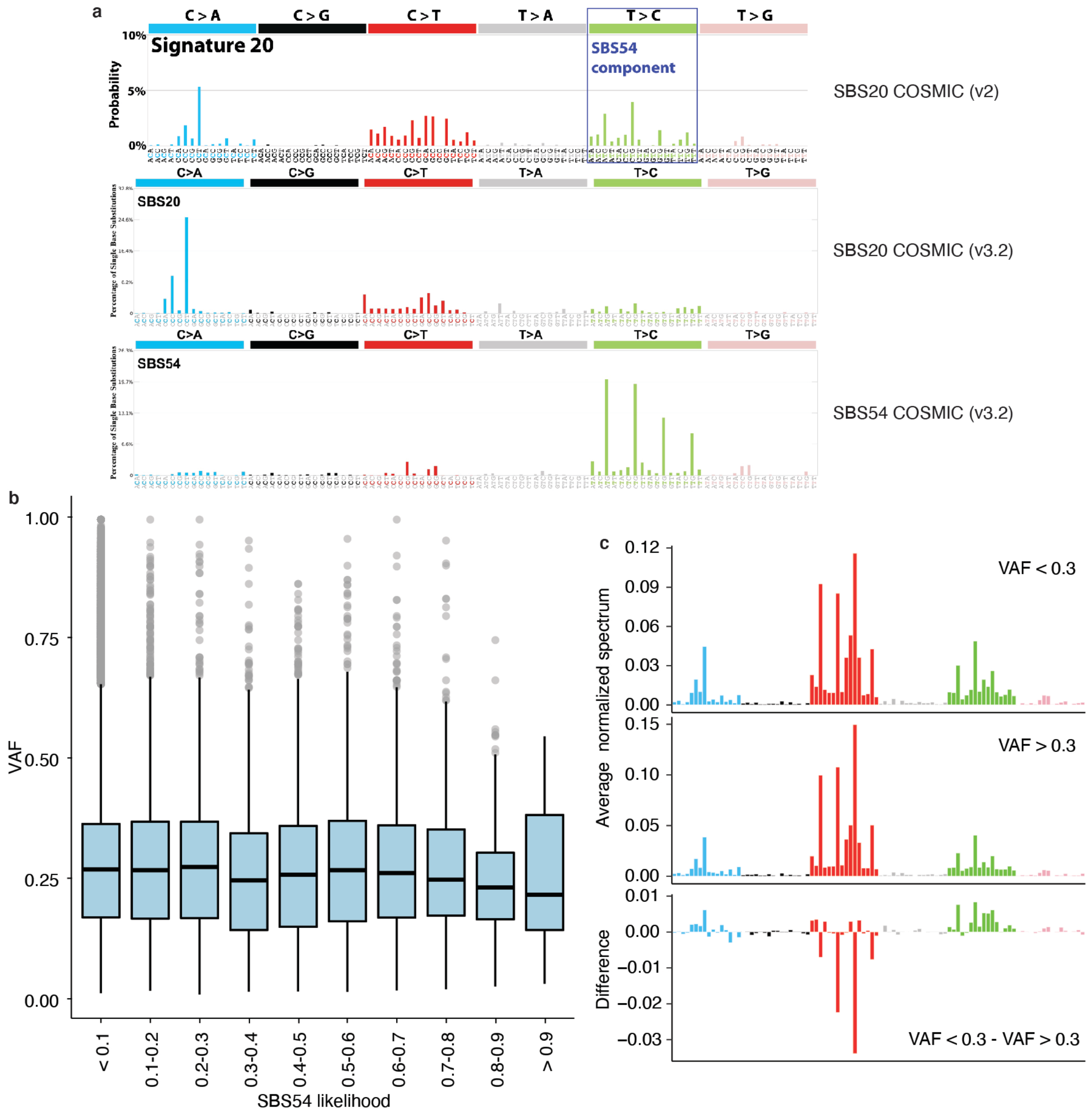


**Supplementary Figure 16. SBS54 high tumors the difference in mutational spectra based on VAF. a.** SBS20 distribution downloaded from COSMIC v2 database, where SBS54 component is marked with a square (Top), the updated SBS20 and updated SBS54 in COSMIC v3.2, respectively for middle and bottom panels. **b.** Box-plot showing variant allelic fractions (VAFs) of mutations in bins of SBS54 likelihood in tumors with non-zero SBS54 exposure in TCGA data. High likelihood means that the mutations are likely to be generated by SBS54, low likelihood means they are likely generated by other mutational processes found in the same tumors. **c.** Comparison of low (<0.3) and high (> 0.3) VAF mutations in SBS54-high TCGA tumors, in top and middle panels respectively. The spectra are normalized to unity and then averaged across samples. Bottom panel show the difference of the two distributions.

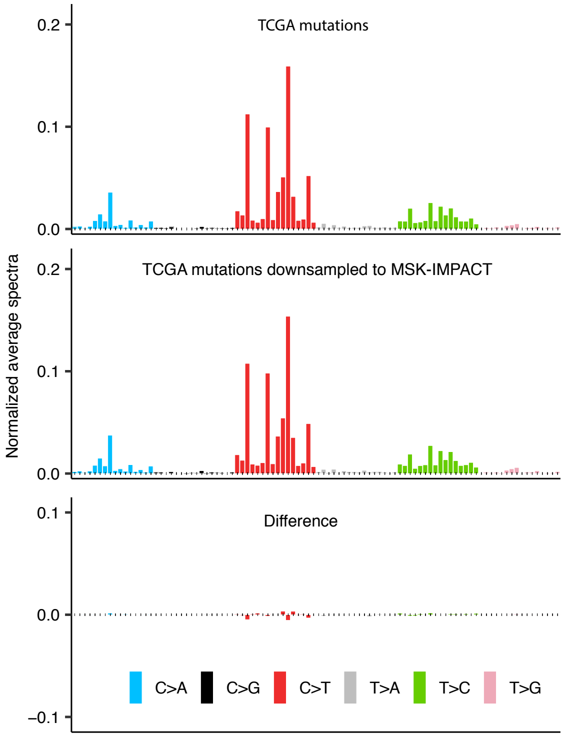


**Supplementary Figure 17. Comparison of all mutations in TCGA data and those downsampled to panels.** The normalized mutational spectra of all the mutations in the TCGA MMRD tumors (top) are compared to the mutations downsampled to panel coverage (middle). Difference of the two spectra is shown at the bottom panel.

Combined analysis of discovery and validation cohort samples
A subset of samples have with PFS (n = 66) and response information (n=65)
A subset has OS information (n = 81) among which part of the samples have MSISensor scores (n = 62)

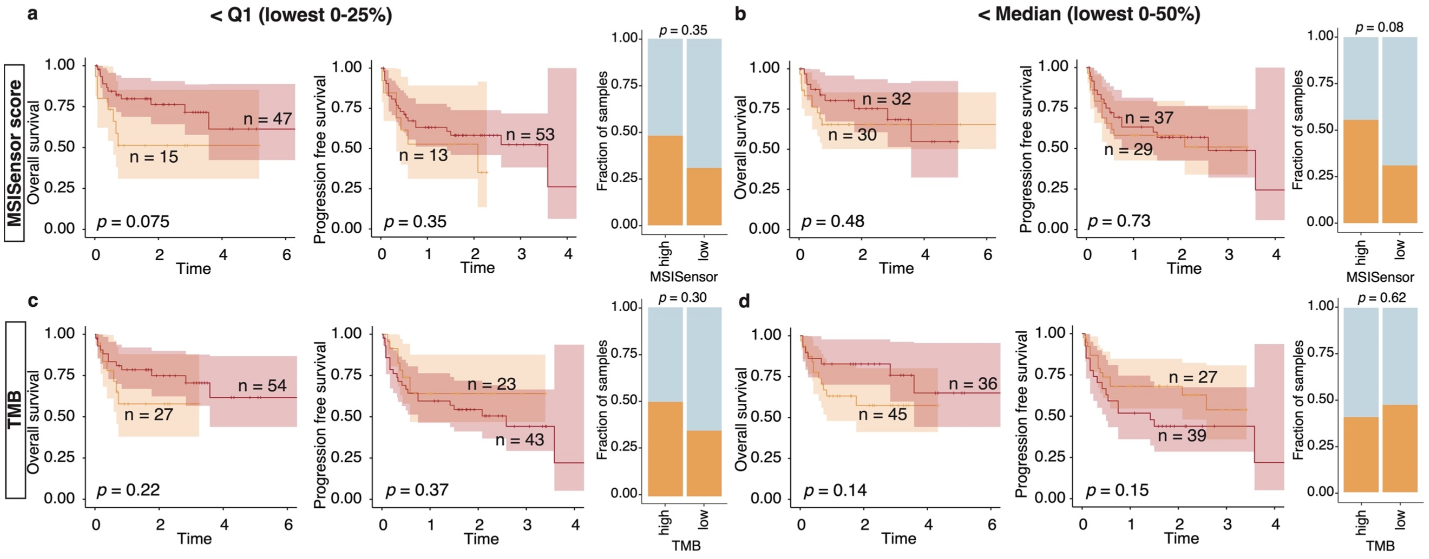


**Supplementary Figure 18. Combined analysis of discovery and validation cohorts for MSISensor and TMB-based selection.** In each panel overall survival, progression free survival and response rate is shown comparing **a-b.** MSISensor high versus low samples stratified based on the first quartile, Q1 **(a)**, and median **(b)**. **c-d.** TMB (per Mb) high versus low stratified based on Q1 **(c)** and median **(d)**. MSISensor Q1: 23.04, median 32.02; TMB Q1: 38.2, median: 48.2 per Mb.

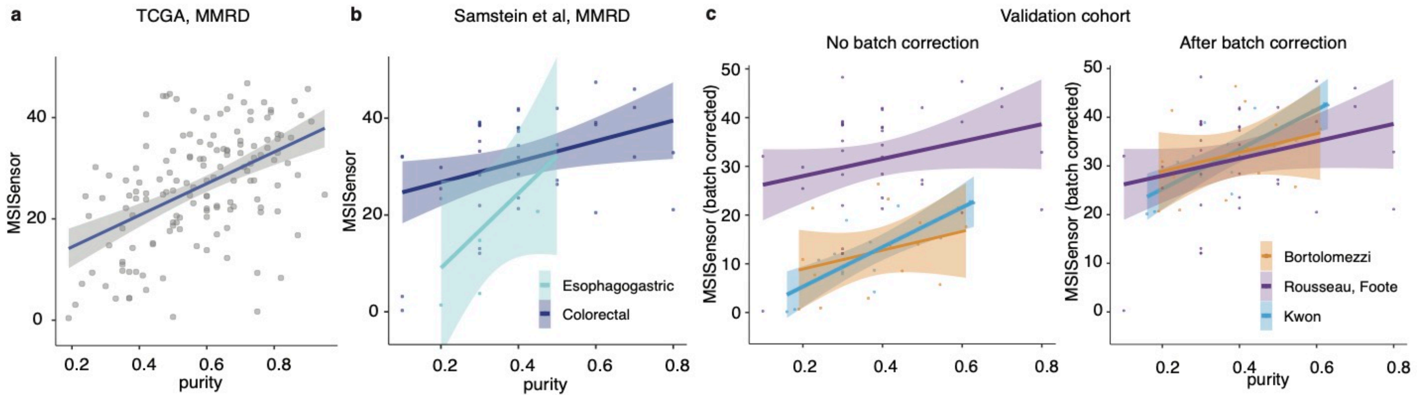


**Supplementary Figure 19. MSISensor-based selection. a.** Dependence of MSISensor score on purity in the TCGA MMRD samples **b.** Same as a but Samstein *et al.* cohort for colorectal and esophagogastric cancers separately. **c.** Same as b but in Bortolomeazzi *et al.*, Kwon *et al.* and Rousseau, Foote *et al.* cohort (left) and after batch correction (MSISensor + 18 for Bortolomeazzi *et al.* and Kwon *et al.*).

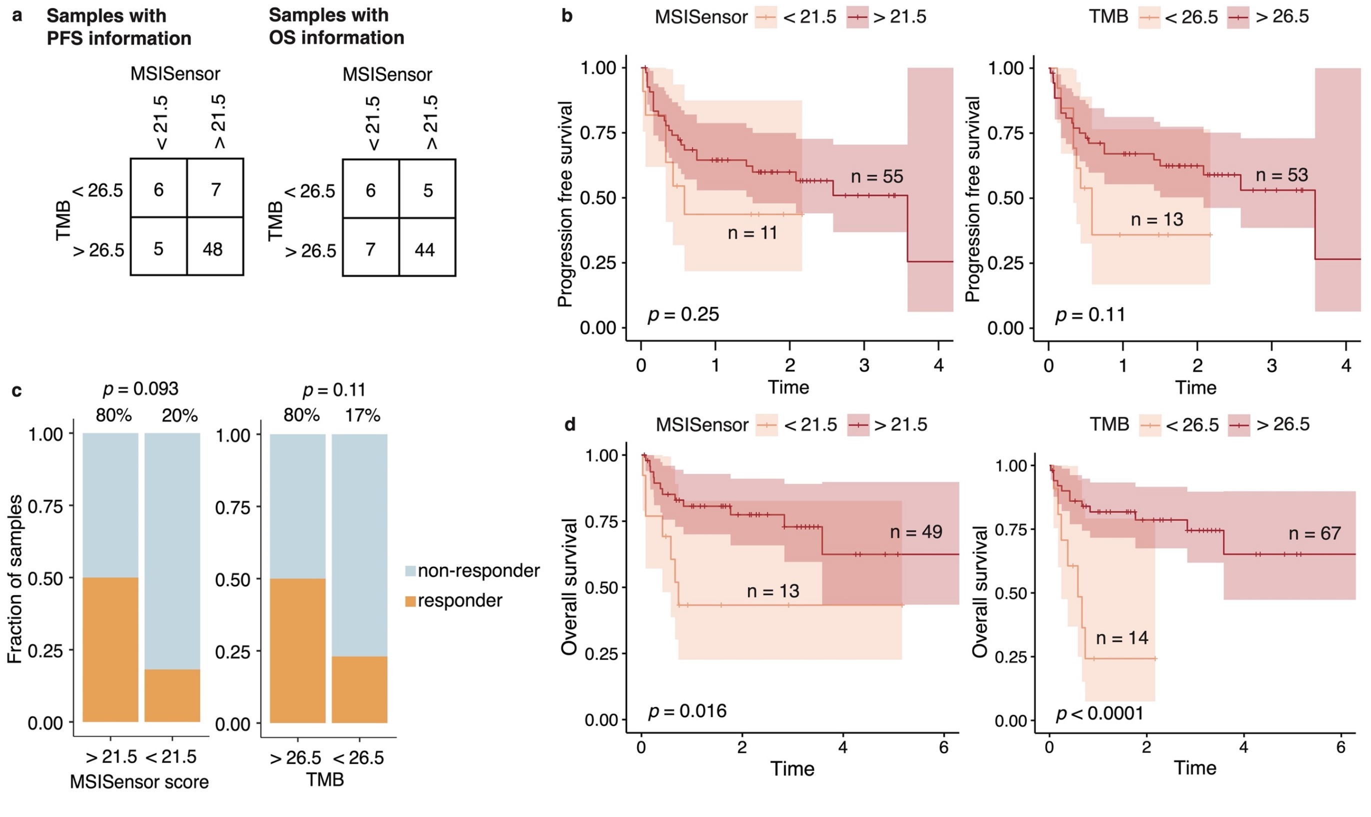


**Supplementary Figure 20. Combined analysis of discovery and validation cohorts for MSISensor and TMB based selection used in previous publications (Kwon *et al.* and Mandal *et al.*). a.** Counts of samples comparing MSISensor and TMB (per Mb) based selections used by two publications from Mandal *et al.* and Kwon *et al.*, respectively**. b.** KM curves for PFS **c.** Response rates and **d.** KM curves for OS, for the same selection.

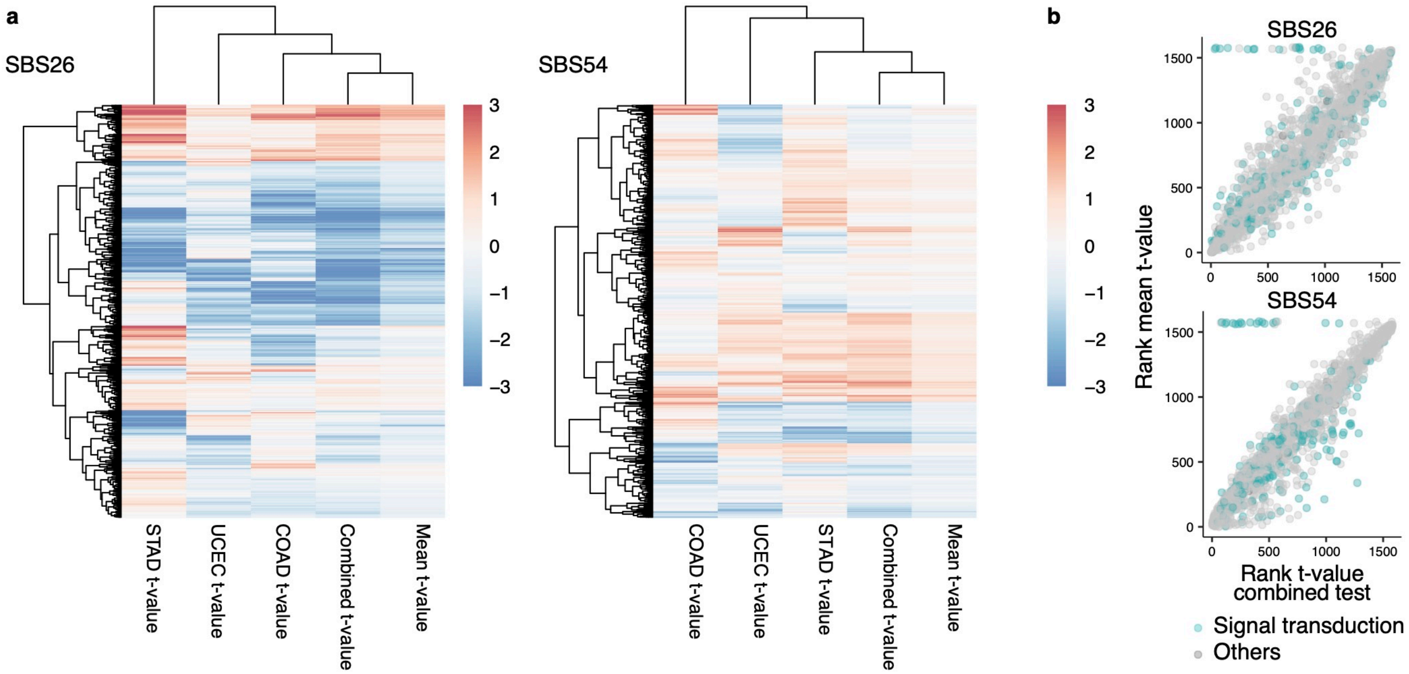


**Supplementary Figure 21. Pathway expression analysis tumor-type specificity. a.** The t-values (truncated at -3 and 3) are obtained from the comparison of SBS26-high and –low (left) and SBS54-high and-low tumors (right) with a t-test. The calculation from specific tumor types (COAD, STAD, UCEC) and their mean value are compared to the calculation with that using the three tumor types together. **b.** The rank of the t-value from the combined calculation are compared to the rank of mean t-value. Few signal transduction pathways were most effected (indicated by marker color), these pathways mainly included FGFR signaling and few others.

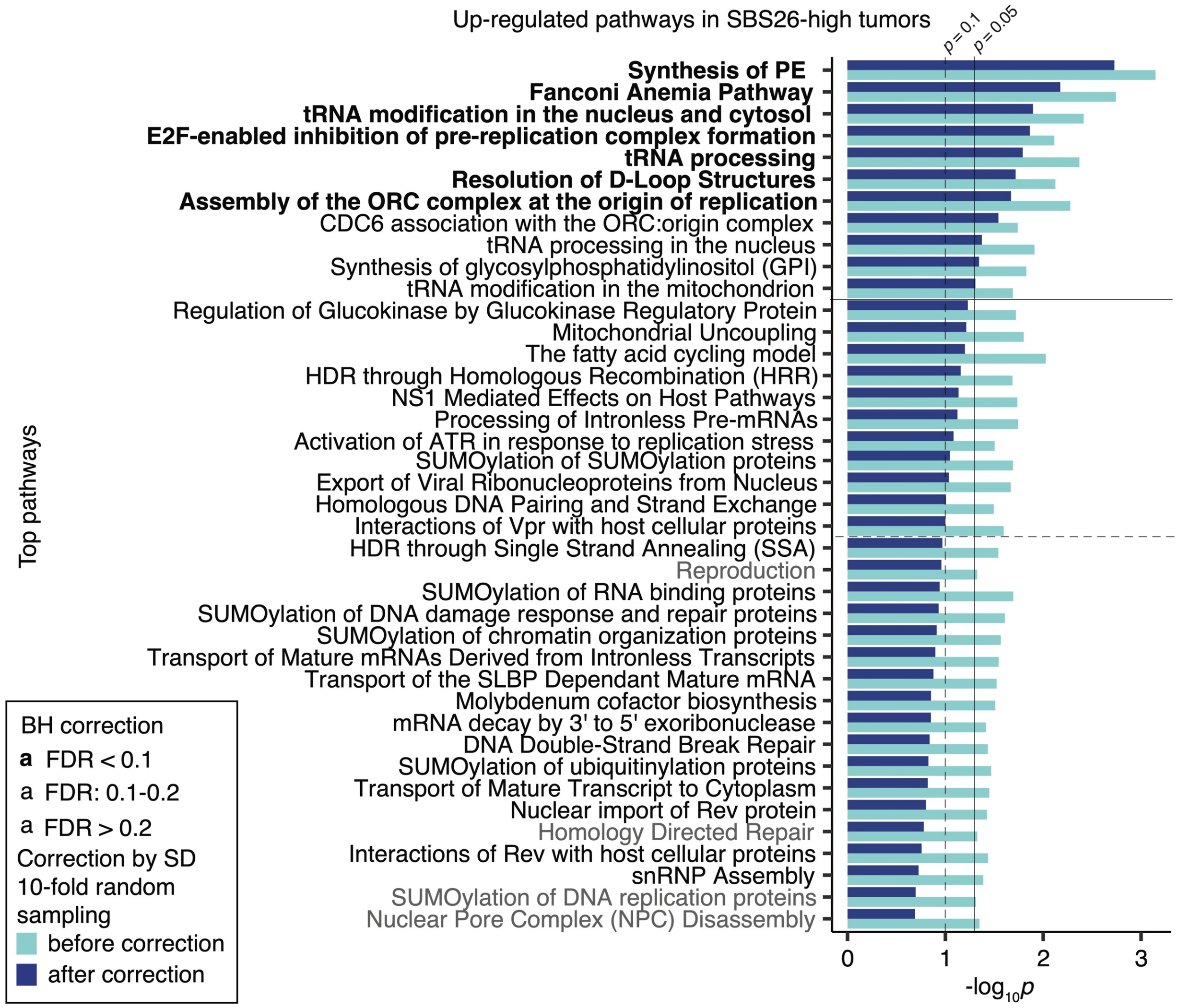


**Supplementary Figure 22. Multiple hypothesis testing correction.** The $-\log_{10}p$ calculated with t-test for pathways with significant upregulation in combined analysis of COAD, STAD and UCEC samples. The values before multiple hypothesis correction in SBS26-high tumors are shown with light blue bars and dark blue after simulation-based correction. The color of the pathway names indicates the BH correction. The vertical and horizontal lines are at $p$ = 0.05, and $p$ = 0.1.

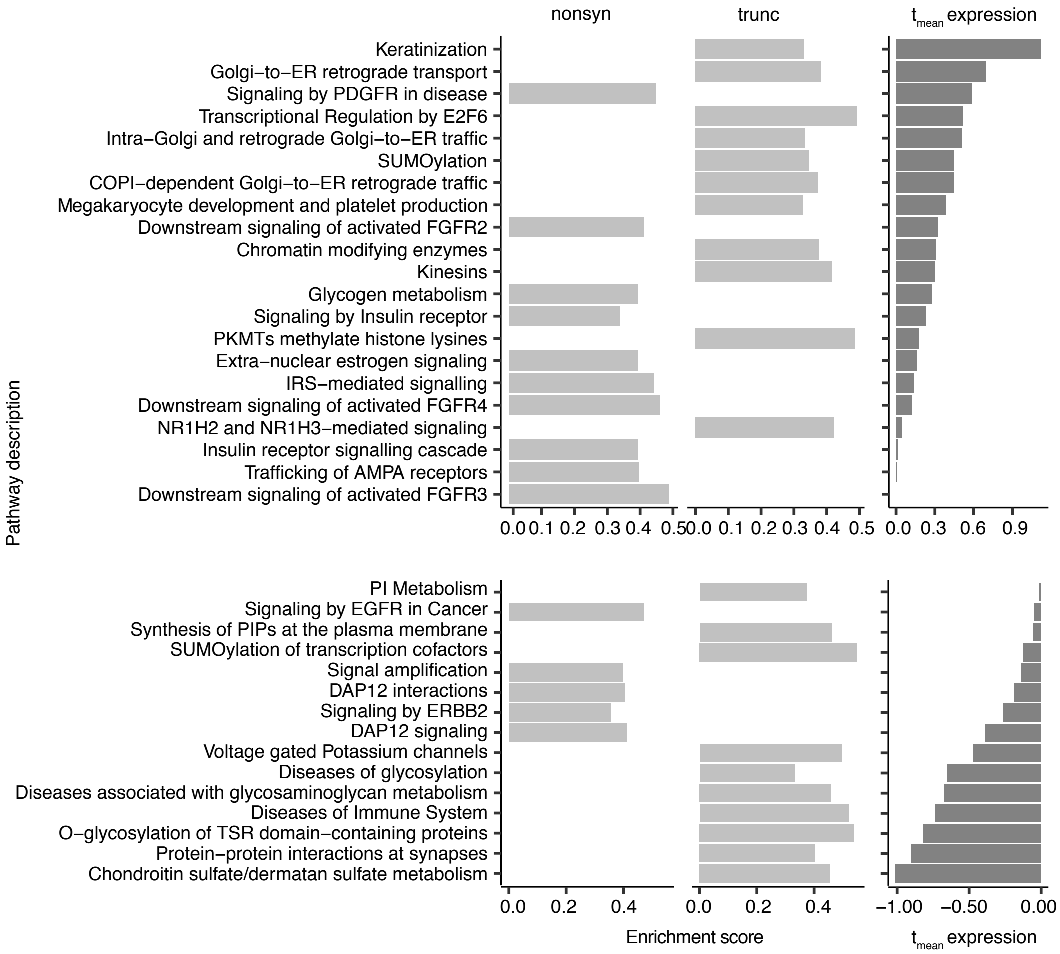


**Extra. Pathways with enriched mutation rate in SBS54-high tumors** (Left three panels) Enrichment scores for pathways with significant enrichment in mutation frequencies ($p < 0.05$) in SBS54-high compared to SBS26-low tumors. (Right) Mean t-values (average of t-values from colorectal, endometrial and gastric cancers) for pathway expression level comparison.
