## Additional_Methods for "Predicting response to immune checkpoint blockade therapy among mismatch repair-deficient patients using mutational signatures"

Gulhan et al.

### 1. MMRD detection with SigMA algorithm

The classifier in SigMA is a gradient boosting classifier that uses features related to microsatellite instability and single base substitution (SBS) signatures for a high precision mismatch repair deficiency (MMRD) detection. The workflow of the algorithm is depicted in Supplementary Figure 1a. The whole-exome sequenced tumors from the TCGA dataset are used as a reference dataset. From this dataset, both the truth tags for MMRD status, which are later used in the training of the machine learning classifiers, are determined and used to simulate targeted gene panels. From the simulated panels, multiple features are extracted, which are used to train gradient boosting classifiers.

This is a similar approach to our previous publication on the detection of the SBS signature homologous recombination deficiency (HRD) from targeted gene panels[1] with some differences. The training is not done to predict the presence of a specific SBS signature but to predict the presence of MMRD. The features are also not limited to those constructed from SBSs. In addition, the reference dataset used in simulations of targeted gene panels and assignment of truth tags are determined from whole-exome sequenced TCGA data and not from WGS datasets because, unlike the HRD tumors, MMRD tumors have hypermutations WES data contains a sufficient number of SBSs for precise signature analysis.

#### a. Validation of the MMRD detection

The MMRD classification with SigMA is compared to MSISensor-based classification (MSISensor score > 10[2]) and to MMR gene immunohistochemistry (Supplementary Figure 1b). The accuracy with respect to IHC and MSISensor scores was 98.7%. The samples that were identified as MMRD and had negative IHC had high MSISensor scores (29.2, 15.0, 5.13), and those identified as MMRP with SigMA but had positive IHC had low MSISensor scores (0.00, 0.00, 0.00, 1.21, 1.07, 1.41, 0.39).

#### b. Features used in gradient boosting classifier

One of the novelties in this particular application of SigMA for MMRD detection is that the classifier is not solely based on SBS signature-related features. It includes variables related to microsatellite instability (MSI) features as well.

MSI-related features can either be:

- Calculated using methods dedicated to identifying mutations at repeat regions such as MSISensor[3] and provided as an input to the SigMA algorithm,
- Alternatively, a proxy can be obtained using indels detected by standard somatic mutation calling algorithms. Indels are overlapped with microsatellite loci, and the count of insertions and deletions overlapping with these areas are used as features. The microsatellite loci are taken as the union set of the repeat regions identified with MSIProfiler[4] and MSISensor[3] algorithms across the whole genome. The MMRD tumors in the TCGA dataset had 80.7% of insertions and 84.3% of deletions in the repeat regions. The two approaches give consistent results (Supplementary Figure 1d)

Hypermutation-related features derived from SBSs are:

- MMRD signature likelihood. The observed mutations are used to calculate multinomial probabilities with respect to the probability distributions defined by the average mutational spectra in clusters of MMRD tumors and POLE-exo mutant tumors. Clusters for MMRD tumors are shown in Supplementary Figure 14. The columns are named in the SigMA output as Signature_X_ml. These distributions are built in the SigMA algorithm for both MMRD tumors and MMRP tumors across 33 tumor types.
- *Cosine similarities* of MMRD SBS signatures. Cosine similarity between sample spectra and distribution of SBS6, SBS15, SBS20, SBS21, SBS26, and SBS54, and POLE SBS signatures (SBS10a, SBS10b) are calculated. Note that SBS6 is not the same as in COSMIC catalog v3.2 (See Online Methods and next sections). Columns are named *Signature_X_c*.
- Signature exposures of MMRD SBS signatures. The exposures are calculated with an iterative NNLS using MMRD signatures together with the signatures found in each tumor type. The tumor type-specific signatures are defined by the signatures identified with non-negative matrix factorization (NMF) from WGS data as discussed in our previous publication[3]. They can be found in *exps_all_msi, sigs_all_msi* columns.
- The likelihood ratios for a unique decomposition. The likelihoods of the observed mutations with respect to the distribution defined by the original decomposition are compared to the likelihoods calculated with an alternative decomposition excluding the signatures with non-zero exposures one by one and using the rest of the signatures specific to that tumor type.

Additional features: counts of SBSs, small insertion and deletions.

#### c. Simulations

Targeted gene panels are simulated from the TCGA dataset. To simulate SBSs and indels, mutation calls were downsampled depending on whether they fall within the coverage of the panel. Targeted gene panels are often with the addition of hotspot microsatellite (MS) loci to improve MMRD detection, and panels of similar library size can have different numbers of MSI loci. As an example, MSI detection in Foundation One CDx panel is done using 114 loci with adequate coverage out of 1897 loci[5], while MSK-IMPACT panels use all of the over 1000 loci for MMRD detection[2].

To provide flexibility in the design of the panels, the MSISensor scores are simulated independently. To see what would be practical values to use to demonstrate the performance of our method, we generated simulations scanning over different numbers of hotpot MSI loci ranging from 10 to 500. MSISensor score refers to the fraction of loci with at least 20 supporting reads that have a loop expansion and contraction. We use the MSISensor scores for the TCGA dataset calculated by the PanCanAtlas group[6] as the input probability and generate a Monte Carlo simulation for a given number of loci, assuming a similar likelihood of mutation at each locus. Based on our simulations, sensitivity as a function of the loci does not improve substantially for values above 100 MS loci (Supplementary Figure 1e). To simulate panels of different sizes, we used the set of genes included in various existing gene panels (see below). Although most of these panels are currently used for detection of germline variants they provide plausible gene selections for panel designs. See below:

| **Panel name** | **Number of genes** | **Size (bp)** | **Size of microsatellite regions (bp)** | **Fraction of microsatellite regions** |
|---|---|---|---|---|
| IMPACT_v2 | > 410 | 1858070 | 105754 | 5.7 |
| AmpliSeq_Illumina_Comprehensive | 409 | 1348636 | 72524 | 5.4 |

| | | | | |
|---|---|---|---|---|
| Illumina_TruSight_170 | 148 | 423539 | 24027 | 5.7 |
| InvitaeMulti-CancerPanel | 82 | 224425 | 12329 | 5.5 |
| Illumina_HotSpot | 50 | 140091 | 7789 | 5.6 |
| InvitaeGynCancerPanel | 23 | 85128 | 4492 | 5.3 |
| InvitaeColorectalCancerPanel | 20 | 53700 | 2934 | 5.4 |

#### d. Gradient boosting classifier (GBC) training

We trained GBCs using the *gbm R* package. The optimal number of trees that minimizes overfitting is selected using 5-fold cross-validation. For the final classifier, all the samples are included in the training. A multi-class training is performed, and samples are categorized into three classes: MMRD, MMRP, and POLE-exo mutant tumors.

#### e. Performance of MMRD detection in simulated panels.

The improvement machine learning-based approach brings, makes it is possible to do accurate MMRD detection down to panel sizes as small as 0.5 Mb. The performance of the GBC is shown in (Supplementary Figure 1c) as dashed and solid black curves for 20 and 100 MS loci, respectively.  Below this value, sensitivity at a 0.3% false positive rate drops substantially. The minimum suggested panel size with previously available computational methods was 1Mb which we are able to expand towards lower panel sizes with our method[7]. The combined score is more effective than each one of the features and more so for smaller panels.

GBC classifier provides improved detection performance compared to each individual feature used in the analysis, including the MSISensor score. In Supplementary Figure 1c, sensitivity for 20 and 100 loci with MSISensor alone are shown as vertical lines and can serve as a baseline of comparison to the GBC model's sensitivity for small and large panels, yielding a 10-15% increase in sensitivity. Note that this estimate is for pan-cancer analysis; when restricted to endometrial, colorectal, and esophagogastric cancers, the increase in sensitivity of detection was found to be smaller, approximately 5%.

### 2. De-novo WGS/WES MMRD signature analysis

We carried out mutational signature analysis using non-negative least squares (NNLS) implementation in the SigProfiler MATLAB package. The MMRD tumors from TCGA WES data are combined with the 40 MMRD WGS samples from the PCAWG dataset. Signatures are matched to the catalog, and if a signature with a good agreement is found, they are replaced by their catalog equivalent. Once the signature distributions are fixed, the exposures are assigned to each sample with non-negative least squares algorithm restricted signatures discovered in MMRD samples and signatures specific to that tumor type. Most of the signatures we discovered were matched to the MMRD or POLE/D1-exo specific mutational signature catalog listed in the signature catalog (COSMIC v3), SBS10a, 10b, 14, 15, 20, 21 and 26.

One of the mutational signatures that has a major contribution to the overall SNV burden in MMRD samples that we discovered was a breakdown of SBS6 into the new distribution and SBS1 (Supplementary Figure 15). A signature that resembles the SBS6 was only discovered when we omit the normalization of mutational spectra of samples before the NMF calculation, meaning that the discovery of the SBS6 mixed with SBS1 is likely to be due to some samples with a high number of SNVS. We refer to this new distribution as SBS6, the refitting replacing the SBS6 in the COSMIC catalog (v3.2).

In addition, we found a mutational signature with NTG > C type mutations, which was previously a part of signature 20 (Supplementary Figure 16a) in the COSMIC catalog (v2), and is currently represented by a new signature in the COSMIC catalog (v3.2) and named SBS54 (distribution shown in Supplementary Figure 15). This signature is annotated as germline contamination, but the discovery of this signature by NMF from the somatic mutations of MMRD samples shows that it is not specific to germline mutations in MMRD tumors. COSMIC catalog (v3.2) is based on analysis of the WGS PCAWG dataset, which may result in the lack of discovery of this signature from somatic mutations considering the smaller number of MMRD samples and the because relative contribution of SBS54 in the genome is smaller than in the coding region of the genome. To further validate its somatic origin, we compared the variant allelic fraction (VAF) distribution in bins of SBS54 likelihood (Supplementary Figure 16b) and observed no increasing trend, which would have been indicative of germline contamination. In another more direct comparison, we selected mutations with VAF < 0.3 and compared their mutational spectrum to the rest of the mutations (VAF > 0.3) in the TCGA dataset (Supplementary Figure 16c). The prior selection would have reduced the contamination from germline variants if there were such issues in the MC3 consensus somatic mutation calls from TCGA data. However, we find that SBS54 is not reduced in the low VAF group. On the contrary, SBS54 is somewhat enriched, which supports the somatic origin of the mutations by SBS54 in MMRD tumors. Note that mutational signatures can generate both somatic and germline mutations. Based on a recent study[8], several signatures discovered germline mutations are very similar to signatures in the COSMIC catalog (e.g., SBS30, SBS8, SBS1, and others). Considering all these points, we are confident that SBS54 in MMRD samples are not originating from germline contamination.

Two additional mutational signatures were found and had small exposures across the MMRD samples. These signatures did not match well with any of the COSMIC signatures. One resembles SBS30 (KL divergence: cos similarity), and the other SBS85. Signatures discovered *de novo* are used for assignment if a good matching to the catalog was not possible. We call these signatures SBS30b and SBS85b. The distribution of SBS30b is similar to the one reported in *UNG* knockout samples, rather than the SBS30 found in tumors with *NTHL1* loss[9,10]. SBS85 was similar to mutational signatures of *POLH*[11].

### 3. Characterization of MMRD samples in the TCGA dataset

We identified 413 MMRD cases, calculated exposures of the SBS signatures, and determined the underlying MMR gene dysfunctions (Supplementary Figure 14).

#### a. Identification of MMRD cases

An initial set of MMRD tumors are selected based on high exposure of SBS6, SBS15, SBS20, SBS21, SBS26, and SBS54 calculated with non-negative least squares (NNLS) algorithm. An initial gradient boosting classifier is trained. Using this classifier, the dataset is expanded, capturing more samples in a second iteration of the calculation. In the training of GBCs the procedure described in Section 1d was followed.

#### b. Signature assignments

The signatures were assigned using NNLS implementation in SigMA, and tumors that carry small exposures of signatures were cleaned for potential false positives using likelihood ratios. Tumors where the exposure of a signature is smaller than 10% and also the likelihood ratio was smaller than 0.65 (Signature_X_l_rat < 0.65) were set to zero.

#### c. MMR gene dysfunction assignments

We define an MMR gene dysfunction if either a reduced expression of 1.5 standard deviations (SD) or a truncating mutation accompanied by a 1 SD decrease in expression. When expression information was not available, we considered genes with a truncating mutation to be MMRD (8 samples). This definition was further refined in order to account for: (1) the fact that the expression of an MMR gene that is not dysfunction can get reduced indirectly by a loss of another MMR gene due to transcriptional co-regulations. (2) The smaller reduction in gene expression by gene dysfunction in samples with lower purity. We selected loose thresholds of 1.5 SD and 1 SD to be able to capture the dysfunction in lower purity samples. Then to compensate for the flexible requirement, we removed cases with a possible false association by looking at the relative reduction in gene expression compared to other MMR genes. In order to avoid a large number of associations per sample, we require the normalized expression of the mutated gene to be reduced by at least 50% as much as the MMR gene with the lowest expression. For example, suppose there is a 3 SD decrease in one gene and only a 1.5 SD reduction in another gene. In that case, we do not consider the latter gene to be dysfunctional unless the gene carries a subclonal truncating mutation. Next, for those cases, where other genes have lower expression than the one with the mutation and 1 SD decrease in the expression, the genes with lower expression than the mutated gene are also defined as dysfunctional.

After this matching procedure, 70% of the samples had a single MMR gene association, and 19% had two. A considerable fraction of samples with two gene associations carried MSH3 and MLH1 dysfunction simultaneously. Some of these samples had frame-shift mutations (K383Rfs*32, N1020Ifs*40, K383Gfs*20) on MSH3 genes, which are likely secondary hits after the initial development of MMRD due to MLH1 dysfunction. Five percent had three dysfunctions, and only 1% of MMRD samples had four gene associations - mostly in low purity samples with high tumor mutation load. Six percent of samples did not associate with any dysfunction. Among samples with POLE or POLD1-exo domain mutations and MMRD, as indicated by the accompanying SBS14, we could not identify the underlying MMR gene dysfunctions for 10 out of 33 cases. The difficulty in determining the dysfunctional gene can be due to a high rate of mutational background. It is also possible that for these samples, the MMRD is subclonal, making it more difficult to pinpoint the gene by expression reduction or mutation. For the rest of the MMRD samples without prominent SBS14, only 4% (16 out of 374) did not have a deficiency in one of the eight MMR genes. For five of these cases, gene expression information was missing, and there was no mutation on the MMR genes we considered. Three of them had truncating mutations on one of the 8 MMR genes but had less than 1 SD reduction in the expression. One sample had POLE3 mutation accompanied by a decrease in its expression, and one sample with POLE4 mutation, and 1 SD reduction in expression, one sample with 2 SD reduction in the expression of POLE4. For 7 cases, we found a moderate (< 1SD) decrease in expression for MGMT, RFC3, RFC5, RPA1, or RPA4. It is also possible that protein level defects may have caused the deficiency[12].

### 4. Differential enrichment analyses in the TCGA data

#### a. Enrichment analysis

*Expression:*
The geneset enrichment scores for the Hallmark and Reactome genesets obtained from MSigDB[13] were calculated with the GSVA package using the following settings using `gsva` function (with parameters `method = 'ssgsea',ssgsea.norm = T, tau = 0.25, mx.diff = F`)

The per-sample GSE scores were compared between SBS26-high *vs* –low and SBS54-high *vs* –low samples with t-tests. We calculated t-values for COAD, STAD and UCEC separately and also for the combination of these three tumor-types. In results shown in Figure 2 and Supplementary Figure 9, we averaged the t-values in COAD, STAD and UCEC in order to account for tumor type specific differences. The multiple hypothesis testing corrections were applied to the pan-cancer comparison instead.

*Mutations:*
For the enrichment analysis, first for each gene we calculated fraction of samples of different mutation types (nonsynonymous mutations, truncating mutations and CN-loss) in SBS26-high and –low or SBS54-high and –low groups. Then using the difference in the frequencies and gsePathway function (with parameters `pAdjustMethod="BH"` and `minGSSize=20`) in the ReactomePA[14] package we calculated the enrichment scores and associated p-values.

#### b. Multiple hypothesis testing corrections

We applied multiple hypothesis testing corrections to the *p*-values obtained in the comparison of SBS26-high versus –low samples (relevant columns were added to Supplementary Table 7) in pan-cancer data. These corrected *p*-values aid in the interpretation of mean t-values reported in Figure 2, and Supplementary Figure 9 (t-tests were performed separately in COAD, READ, and UCEC, separately and t-values are averaged). We followed two approaches for the correction:

- We used the Benjamini-Hochberg (BH) procedure which controls the false discovery rate (FDR) instead of the familywise error rate. With 20% FDR, almost all the pathways (93%) with a significant ($p < 0.05$) differential expression in SBS26-high tumors before multiple hypotheses testing corrections were captured, among which Fanconi Anemia and DSB repair pathways were found (Supplementary Figure 22; showing only the upregulated pathways, see Supplementary Table 7 for all pathways).
- BH procedure is still likely to cause false negatives because genesets defining different pathways are nested, resulting in correlations between GSE scores. Therefore, as an alternative, we followed a non-canonical approach. We performed 10-fold simulations with random sampling and rerunning the full workflow of gene set enrichment analysis. We calculated the standard deviation of the $–\log_{10}p$ of the simulated data and subtracted it from the $-\log_{10}p$ obtained in our data.

For SBS54-high tumors, BH correction yields results with only FDR > 20%. Simulation-based correction still yields some pathways with $p < 0.05$ (MYC targets, base excision repair pathways among the top and with $p < 0.1$).
